# Effect of Antiseptic Mouthwash/Gargling Solutions on SARS-CoV-2 Viral Load: A Randomized Clinical Trial

**DOI:** 10.64898/2026.05.20.26353686

**Authors:** Sepideh Banava, Allan Radaic, Pachiyappan Kamarajan, Nancy F. Cheng, Yvonne L. Hernandez-Kapila, Stuart A. Gansky

## Abstract

**Background:** The COVID-19 pandemic has caused significant global mortality. Despite declining infection rates, new variants of SARS-CoV-2 continue to emerge, necessitating new prevention strategies.

**Objective:** This study aimed to evaluate the effect of four over-the-counter (OTC) antiseptic mouthwash/gargling solutions in the U.S., compared with a distilled water control, on SARS-CoV-2 viral load across multiple oral and oropharyngeal sample types.

**Methods:** This pilot single-center randomized controlled clinical trial enrolled adults in the San Francisco Bay Area, California, who tested positive for COVID-19. Participants were randomized to distilled water, chlorine dioxide, hydrogen peroxide, cetylpyridinium chloride, and essential oils. Participants were instructed to rinse and gargle four times daily for four weeks using standardized instructions to ensure protocol adherence. Samples were collected on Days 1, 7, and 28 and analyzed using reverse transcription–quantitative polymerase chain reaction (RT-qPCR). The primary outcome was the change in SARS-CoV-2 viral load from baseline to Day 28, assessed using cycle threshold (Ct) values. Secondary outcomes included self-reported clinical symptoms and hospitalization.

**Results:** Forty-nine participants completed the study. No mouthwash demonstrated a statistically significant reduction in SARS-CoV-2 viral load over time. Cetylpyridinium chloride showed a transient increase in Ct values on Day 7 that was not sustained on Day 28. At baseline, throat swab samples had the lowest Ct values across all sample types. Due to limited subgroup sample sizes for secondary outcome measures, no statistical or moderator analyses were conducted.

**Conclusion:** Further large-scale randomized trials are needed before recommending antiseptic mouthwashes for SARS-CoV-2 prevention or management.

**Trial Registration:** ClinicalTrials.gov NCT04409873

## Introduction

The COVID-19 pandemic has resulted in devastating global mortality, with approximately 7.1 million deaths worldwide and over 1.2 million deaths in the United States (US) [1]. Despite declining infection rates, the CDC reports about 200 weekly US deaths, along with persistent long COVID cases [2].

SARS-CoV-2, COVID-19’s causative agent, undergoes frequent mutations, leading to new variants and successive infection waves, necessitating ongoing adaptations in prevention and treatment strategies [3–5]. The primary transmission mode remains respirable particles inhaled into the oronasopharynx, leading to viral colonization in the upper respiratory tract (URT) [4, 5]. While mild cases typically present with symptoms such as fever, cough, and nasal congestion [3], severe cases can progress to pneumonia, acute respiratory distress syndrome (ARDS), and multi-organ failure [6–8]. Individuals with comorbidities such as hypertension, diabetes, and cardiovascular disease remain at higher risk for severe disease, hospitalization, and mortality [9]. Furthermore, certain racial and ethnic minority groups, including American Indian or Alaska Native, Black or African American, and Hispanic or Latino/a individuals, have been disproportionately affected with higher infection rates, hospitalization, and death compared to non-Hispanic White populations [10, 11]. The healthcare system has faced significant challenges throughout the COVID-19 pandemic. Frontline healthcare professionals (HCPs), including dental practitioners, were at an increased infection risk due to frequent exposure to aerosols generated during dental and medical procedures [12]. According to the CDC, in April 2020, among 315,531 COVID-19 cases, 9,282 (19%) were identified among HCPs, which was an underestimation as HCP status was available for only 16% of reported cases nationwide [13]. Dentists as HCPs, at the pandemic onset, faced challenges in ensuring safe dental treatments for both patients and themselves due to the increased risk of occupational exposure to aerosols.

With growing public concern about infection control, interest in mouthwashes as a potential preventive measure surged globally during the pandemic [14]. Mouthwashes have long been used for oral hygiene and dental disease prevention [15]. Studies on SARS, Middle East Respiratory Syndrome (MERS), and Influenza H5N1 have shown that mouthwashes with an antiseptic ingredient (e.g., chlorhexidine gluconate, povidone-iodine, chlorine dioxide, cetylpyridinium chloride, hydrogen peroxide, and even water via physical action) can reduce viral load and transmission [16–18]. At the COVID-19 onset, various antiseptic mouthwashes were suggested to potentially mitigate against SARS-CoV-2 infection [19]; however, given the limited evidence at the time, the American Dental Association (ADA) and CDC recommended that dentists use a pre-procedural mouth rinse containing hydrogen peroxide (1–1.5%) before dental treatment to potentially reduce oral viral load [20]. At the time, evidence for hydrogen peroxide or other antiseptic mouthwashes against SARS-CoV-2 in vivo remained limited and inconsistent.

Subsequent publications have yielded contradictory and inconclusive findings regarding the efficacy of different mouthwash ingredients, concentrations, rinse durations, and sampling methodology to inactivate the virus. Some *in vitro* studies have reported that certain over-the-counter (OTC) mouthwashes can inactivate SARS-CoV-2 within 30 seconds, while others found partial inactivation or transient effects [21, 22]. A German *in vitro* study found that three of eight tested OTC antiseptic mouthwashes, including those with benzalkonium chloride, chlorhexidine, and povidone-iodine, could inactivate SARS-CoV-2 within 30 seconds, while the remaining five had partial effects [22]. Similarly, a US *in vitro* study demonstrated that a CloSYS Ultra-Sensitive oral rinse reduced SARS-CoV-2, SARS-CoV, and Influenza A H3N2 viral loads, with a tenfold greater reduction for SARS-CoV-2 than for SARS-CoV within 30s [23]. Additionally, a clinical trial in Bangladesh found that 1% povidone-iodine (PVP-I) used as a mouthwash, gargle, and eye/nose drop significantly reduced COVID-19-related morbidity, mortality, and hospitalization rates [24].

Despite these promising results, randomized controlled trials are still needed to establish evidence-based recommendations for antiseptic mouthwash use against SARS-CoV-2, including optimal concentration, chemical formulation, and mouthwash use duration. In clinical studies, SARS-CoV-2 viral load is commonly quantified using reverse transcription–quantitative polymerase chain reaction (RT-qPCR) cycle threshold (Ct) values, which are inversely related to viral RNA levels. However, Ct interpretation is influenced by assay detection limits, sampling method, and within-subject variability over time. Analytical approaches that appropriately account for censoring and within-subject correlation are essential for accurately evaluating viral load dynamics in outpatient infections.

This study aimed to evaluate the effects of four OTC antiseptic mouthwash/gargling solutions marketed in the US, compared to a distilled water control, on SARS-CoV-2 viral load in unstimulated saliva, throat wash (gargle lavage), and oropharyngeal/throat swab samples. Specifically, the study objectives were to:

a. Compare the cycle threshold (Ct) values of SARS-CoV-2 in unstimulated saliva and throat wash samples before and after rinsing with the randomly assigned product on Day 1 (baseline), Day 7, and Day 28; b) Evaluate clinical symptom progression and healthcare utilization (e.g., hospitalization); c) Assess whether tobacco use, marijuana smoking, or vaping modify the primary outcome measures; and d) Monitor participants’ health status over 12 months.

The null hypothesis posits that antiseptic mouthwashes will not significantly change Ct values in any sample type over the 28-day study period. Findings from this study will provide critical insights into whether readily available OTC mouthwashes can serve as effective adjunctive measures in reducing SARS-CoV-2 load.

## Materials and Methods

### Study design

This study was a pilot, single-center, 5-arm, parallel-group, double-masked (participant, outcomes assessor), randomized controlled clinical trial conducted between 2021 and 2022 in the San Francisco Bay Area (California, USA) on adults who tested positive for COVID-19 confirmed by RT-qPCR testing. Participant recruitment was conducted from 01/04/2021 to 18/10/2021, and participants completed a 4-week intervention period and were subsequently followed at quarterly intervals for an additional 11 months, with final follow-up completed on 18/10/2022. This clinical trial was approved by the Institutional Review Board at the University of California, San Francisco (UCSF) (IRB number: 20-30874). All participants provided informed consent.

### Sample Size

A total of 150 individuals were planned for this pilot trial to provide precise estimates of effect size for a future definitive trial with at-risk individuals. A sample size of 24 for a mouthwash group and 24 for the distilled water control group was estimated to have 80% power to detect a 2-fold difference in mean values between the 2 groups with a 2-sided alpha=0.05. Assuming 80% retention through 28 days, 30 participants per group at baseline (150 total) were planned for full enrollment. This report presents a pre-planned interim analysis of the first 49 participants who completed the 28-day study, after which the trial was stopped early.

### Participant Recruitment

Table 1 outlines the study inclusion and exclusion criteria. Two recruitment strategies were utilized: 1) Study Team Approach: As part of the UCSF Clinical and Translational Science Institute (CTSI) Participant Recruitment Program (PRP), Dr. Sepideh Banava (SB) reviewed the UCSF COVID-19 positive patient list and their electronic health records (EHR). SB initially triaged the list to extract those 18 years and older with a positive test in the last 7 days. Then, the study team contacted patients via phone, explained the study details, assessed eligibility criteria, and asked about their interest in participating in this clinical trial; 2) Individual Approach: Individuals who tested positive for COVID-19 contacted the study team via contact information on study flyers at the UCSF testing centers and the study’s clinical trial webpage at UCSF.

**Table 1.**
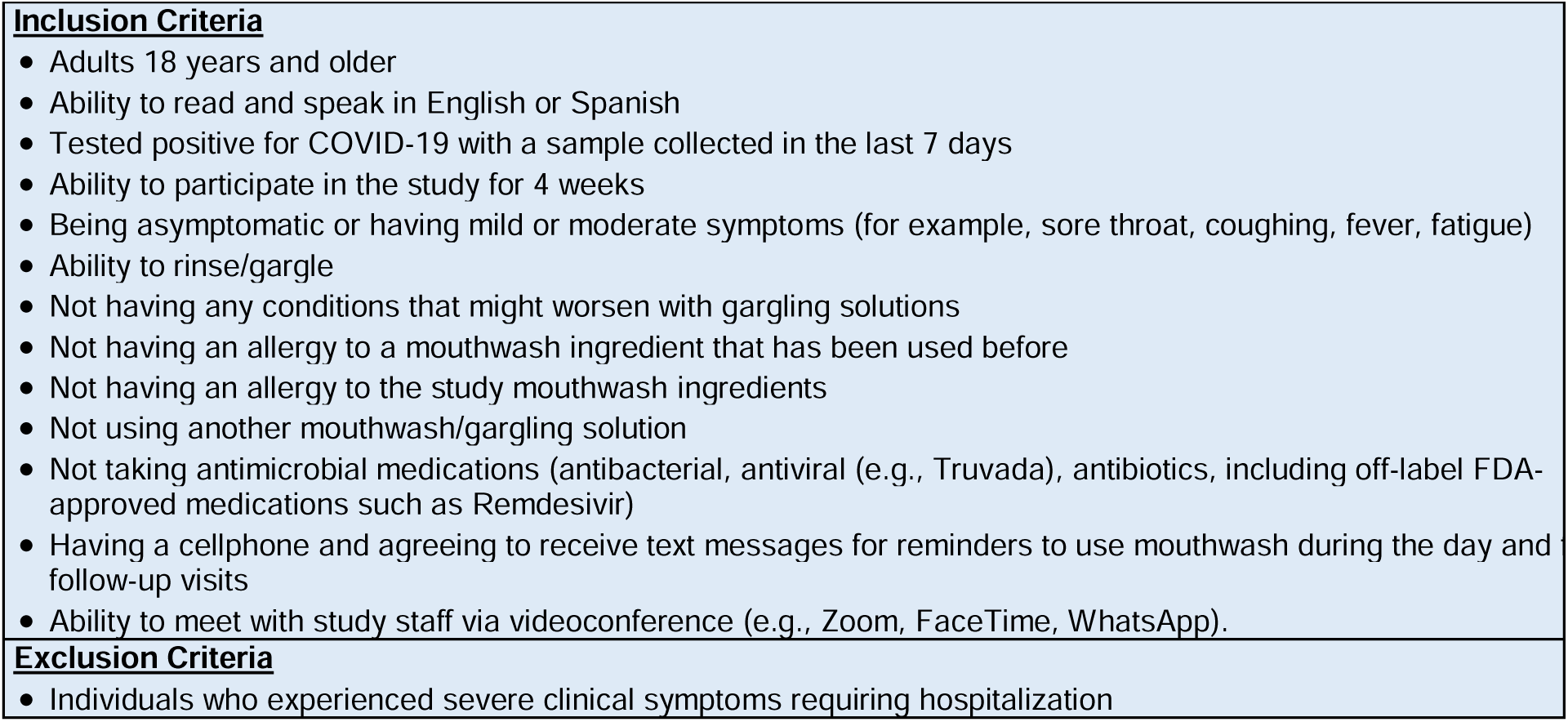
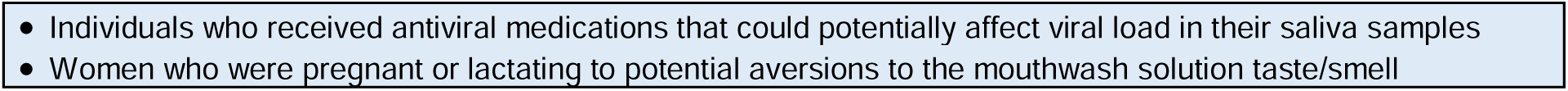
Inclusion and Exclusion Criteria.

Upon contacting participants, participants’ eligibility was assessed and confirmed based on inclusion/exclusion criteria via an electronic eligibility form in the Research Electronic Data Capture (REDCap) hosted at UCSF [25, 26]. REDCap is a secure, web-based platform designed for research data capture, providing an intuitive interface, audit trails, and export to common statistical packages.

After eligibility assessment during a telephone or videoconference call, eligible participants were emailed an IRB-approved electronic informed consent form through REDCap and provided consent electronically prior to enrollment. The form explained the study goals, sample collection procedures, lab tests, and confidentiality of the participants’ information. After signing the consent form, participants received and completed an electronic baseline questionnaire, which included 17 questions about demographics, health conditions, symptoms, vaccination status, and oral hygiene practices (Appendix A).

### Randomization and Masking

After completing the baseline questionnaire, participants were randomized in real-time via REDCap into one of five study groups to receive the assigned mouthwash and corresponding sampling package. The study was double-masked, with both participants and outcome assessors masked to group allocation. To maintain masking, all mouthwash bottles had identifying labels and product descriptions removed or covered prior to distribution. Laboratory personnel performing RT-qPCR assays received samples labeled only with barcode-based participant IDs and remained masked to intervention assignment throughout the study. (However, bottle shapes differed for each product so a participant may have been able to guess his/her assigned product.) Unmasking was planned only if it would be necessary for participant safety.

### Intervention

Participants were randomly assigned to one of five mouthwashes and instructed to rinse their mouths and gargle four times daily (after each meal and before bedtime) for 28 days. The amount (dose) and timing (duration) followed manufacturer recommendations. For the control group the timing was chosen based on previous studies [27, 28].

Table 2 shows details on the study groups, mouthwash ingredients, and dosages. The mouthwash selection criteria were based on previous *in vitro* study results and included being an OTC mouthwash, alcohol-free, gluten-free, fluoride-free, and legally marketed in the US (Table 2).

**Table 2.** Study Groups and Mouthwash Details.

| Table 2. Study Groups and Mouthwash Details |  |  |
| --- | --- | --- |
| Group | Mouthwash | Dosage |
| 1 | Distilled water (Control); Smartwater (original); Glacéau, USA | 20 mL, 60s (30s rinse, 30s gargle) |
| 2 | Chlorine Dioxide; CloSYS Ultra Sensitive; Rowpar Pharmaceutical Inc.; USA | 15 mL, 40s (20s rinse, 20s gargle) |
| 3 | Hydrogen Peroxide (1.5%); Oral-B Mouth Sore; Proctor & Gamble; USA | 20 mL, 60s (30s rinse, 30s gargle) |
| 4 | Cetylpyridinium Chloride (0.07%); Crest Pro-Health Multi-Protection; Proctor & Gamble; USA | 10 mL, 60s (30s rinse, 30s gargle) |
| 5 | Essential Oils (Eucalyptol 0.092%, Menthol 0.042%, Methyl Salicylate 0.060%, Thymol 0.064%). Listerine Zero; Johnson & Johnson; USA | 20 mL, 30s (15s rinse, 15s gargle) |

Participants received their study package on recruitment day, before Day 1 (baseline) sample collection. The package included sample collection kits for Days 1, 7, and 28 with unique barcodes for tracking and masking purposes (known only to SB and the research assistant). It also contained the assigned randomized mouthwash and illustrated sample collection instructions.

### Sample Collection Procedure

Staff explained the sample collection process via videoconference on the morning of Days 1, 7, and 28, to participants immediately after waking and before eating/drinking anything while on videoconference. If participants had specific eating routines, the sample collection was scheduled 60 minutes after their last meal. On recruitment day, Day 1 sample collection was scheduled during an encrypted videoconference. Participants received a text message the day before, reminding them not to eat, drink, or smoke at least 60 minutes before sample collection. The sample collection process for Days 1, 7, and 28 was as follows:

Day 1: at the scheduled videoconference time, SB instructed participants to collect five samples step-by-step in the following order:

a. <u>Throat swab</u>: Participants were instructed to use an oropharyngeal swab (OP) (Zymo Collection Swab, Zymo research, USA) to swab the posterior/pharyngeal wall of their throats in front of a mirror.
b. <u>Pre-rinse unstimulated saliva</u>: Participants were instructed to collect unstimulated saliva (i.e., drooling saliva without any force or spitting) in a pre-marked container.
c. <u>Pre-rinse throat wash (gargle lavage)</u>: After 10 minutes, participants rinsed their mouth and throat with 5 mL of distilled water for 30 seconds and collected the throat wash in a sterile container. SB demonstrated how to prepare the saliva sample transportation kit (i.e., the DNA/RNA shield reagent, Zymo Research, USA). This step was required to preserve the sample, inactivate the virus, and ensure safe transport, thereby reducing the risk of inadvertent contamination.
d. <u>Rinsing with assigned mouthwash</u>: Participants were instructed to use a provided graduated cup to measure the assigned mouthwash and rinse and gargle according to the instructions on the label.
e. <u>Post-rinse unstimulated saliva</u>: Participants were instructed to wait five minutes and then collect post-rinse unstimulated saliva in designated containers.
f. <u>Post-rinse throat wash</u>: Participants were instructed to collect post-rinse throat wash with the provided 5 mL of distilled water vial (Appendix B).

At the end of the session, participants were instructed to prepare the pick-up package for the research staff. The Day 1 sample collection took about 60-75 minutes.

Participants were instructed to continue using the same mouthwash from Day 1 and throughout the study (28 days). The study participants were instructed not to use any other mouthwashes or take any antimicrobial medication (e.g., antibacterial, antiviral, antibiotics including off-label usage such as Remdesivir) during the study. However, they were instructed to continue practicing their usual home oral hygiene care (tooth brushing with toothpaste, and flossing). In case of a pre-scheduled dentist appointment, they were asked to inform their dentists about participation in this trial and mouthwash use should any dental treatment be done.

Starting from Day 1, participants received daily SMS text reminders about mouthwash use, upcoming sample collection sessions (Days 7 and 28), and a link to a brief REDCap daily questionnaire asking about rinse use frequency, duration, method, and any potential adverse effects (e.g., mouth burning, taste change, mouth ulcer or sore, inflammation or swelling, tooth stain, redness). Study staff encouraged participants to contact staff if they encountered any issues with the assigned mouthwash.

Days 7 and 28: On a scheduled 15-minute videoconference, participants were instructed to collect throat wash samples via the provided sample collection kit.

All collected samples were picked up by the research team and delivered to the lab for testing.

### Outcome Measures

The primary study outcome measure was change in SARS-CoV-2 viral load in unstimulated saliva and throat wash samples from baseline to Day 28 via RT-qPCR.

Secondary outcome measures included change in self-reported clinical symptoms, and change in healthcare utilization or hospitalization from baseline to Day 28. Additional outcomes included change in SARS-CoV-2 viral load in tobacco users, marijuana smokers, or vapers.

### Laboratory Procedures

Samples were transported to a Biosafety Level 2 Plus (BSL2+) laboratory following WHO guidelines and stored in a −80°C freezer (Thermo Fisher Scientific, USA) until further processing. The following steps were performed to assess SARS-CoV-2 in collected samples.

<u>i. RNA Extraction-</u> RNA was extracted from both collected saliva and oropharyngeal samples using the Quick DNA/RNA Viral Kit (Zymo Research, USA), following the manufacturer’s instructions. First, viral DNA/RNA buffer was added to 800µl of sample (2:1 v/v). Then, each sample was added to Spin Columns with a 2 mL collection tube and centrifuged for 2min at 12,000 RPM. Next, the flow-through was discarded, and the columns were washed by adding 500µl of the viral wash buffer and centrifuged for 30 seconds at 12,000 RPM twice. Subsequently, 500µL of ethanol (100%) was added to the columns and centrifuged for 1min at 12,000RPM for complete removal of the washing buffer. Then, the purified RNA from each sample was harvested from the columns by adding 50 µL of DNase/RNase-free water to the columns, incubating for 5min at RT, and centrifuging for 1min at 12,000 RPM. Finally, the RNA content in the samples was measured using a Nanodrop One (Thermo Fisher Scientific, USA) and stored at −80°C until further processing.
<u>ii. Reverse Transcription Quantitative Polymerase Chain Reaction (RT-qPCR)-</u> After measuring the RNA content for all the samples, we determined that the least concentrated sample had 1.8ng/µL of total RNA. We used this sample concentration as the reference against which to normalize all other samples, such that the total RNA tested for each sample was 15.12ng of RNA per reaction; and in this case, the maximum sample volume for the reaction was 8µL.

Then, the presence of SARS-CoV-2 viral RNA in the samples was checked by RT-qPCR, using the Luna® Universal One-Step RT-qPCR Kit (New England BioLabs, USA), according to the manufacturer’s instructions. Briefly, 10µL of Luna Master-Mix, 1µL of Enzyme Mix, 0.8µL of forward and reverse primers (10µM) for N1, N2, and Rnase P (RP) genes, 0.4µL (10µM) of their probes (Table 3) (Integrated DNA Technologies, USA), and 15.12ng of sample RNA were added to each well, with each sample being tested in triplicate. Table 3 displays “Forward and Reverse Primers” used in the assay. The probes were 5’-end labeled with either FAM, SUN (equivalent to VIC) or TAMRA and 3’-end quenched with BHQ.

**Table 3.**
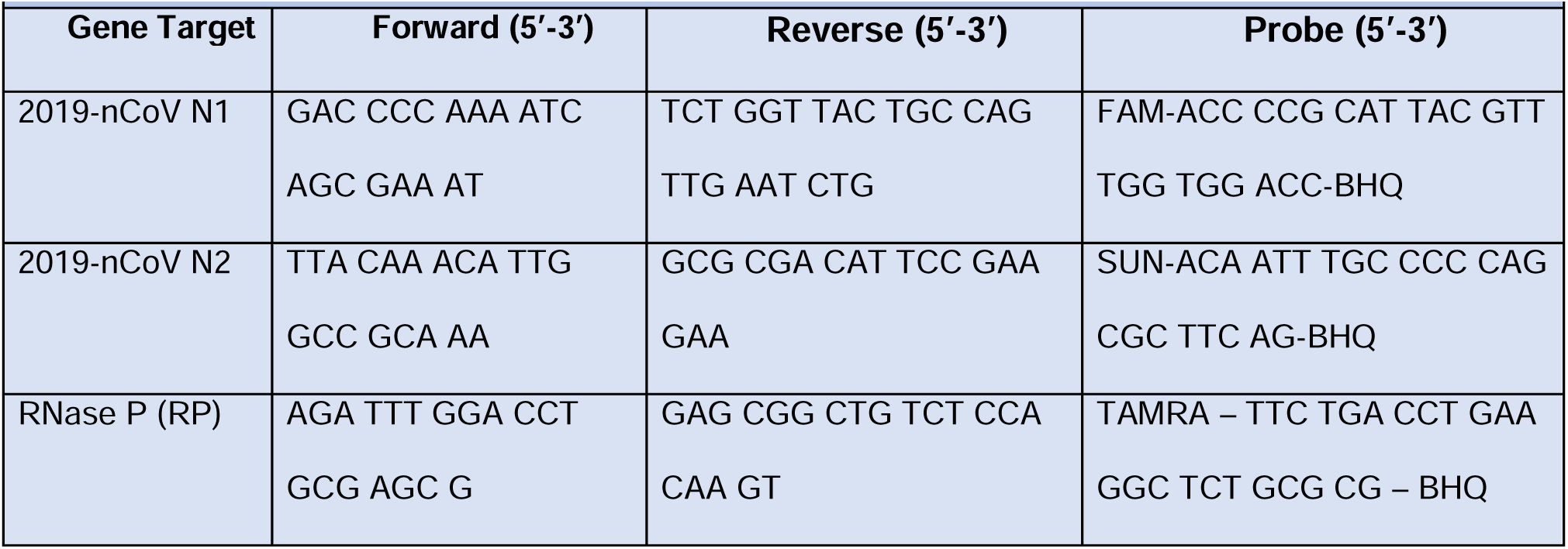
Forward and Reverse Primers Used in the Assay.

In our experimental setup, we amplified 3 different genes in the same reaction - N1, N2, and RP genes – thus, forming a multiplex PCR reaction. Both N1 and N2 are derived from SARS-CoV-2 nucleocapsid protein, whereas RP is derived from human cells. Specifically, amplification of both N1 and N2 are indicative of COVID-19 infection, whereas RP amplification is the clinical sample internal control, indicating a successful RNA extraction and RT-qPCR amplification.

In the assay, we included a negative (no template) control, which consisted of adding RNase-free water in place of the sample to assess if reaction solutions were contaminated with human or SARS-CoV-2 RNA. We also included a SARS-CoV-2 positive control, which was used to assess PCR run integrity. The positive control contained templates of the SARS-CoV-2 complete nucleocapsid gene, including the N1 and N2 gene targets, but not the RP gene. Thus, these controls were used to validate both aspects of the assay.

After placing the samples into their respective wells, the plate was sealed with optically transparent film to avoid evaporation artifacts and centrifuged at 3,000RPM for 1min to remove any bubbles in the solutions. Finally, the plate was read in a QuantStudio 3 Real-Time PCR System (Thermo Fisher Scientific, USA) programed with the thermocycling protocol indicated by the Luna® Universal One-Step RT-qPCR Kit (New England BioLabs, USA) for 40 cycles, and the Ct values from each sample amplification curve were obtained. ROX dye (present in the master mix) was used as a reference to obtain the fluorescence threshold of each reaction.

### Data Collection and Management

Participants’ health information was collected via REDCap questionnaires including eligibility, baseline, and daily and long-term follow-up questionnaires. Email addresses and phone numbers, the only personal identifying information, were kept confidential and saved in REDCap on secured servers and encrypted laptops and deleted when the study ended after the 12-month follow-up. SB regularly supervised data entry and its quality. The RT-qPCR test results of collected biospecimen viral loads were recorded and saved as Ct values.

### Termination of the trial

An interim analysis with analyst masked to was carried out after recruiting the first 50 participants, the estimated sample size for the first stage of the study. During the statistical analysis phase, the funder withdrew the funding, and given the study results, the co–principal investigators decided to stop recruitment and terminate the trial on 09/10/2022; however, the 12-month follow-up of the last recruited participant finished on 18/10/2022.

### Statistical Analysis

Multiple statistical approaches consistent with the *priori* analytic plan were used to evaluate the impact of different mouthwashes on Ct values (inverse viral load) among study groups. Survival analysis using Cox proportional hazards regression (PROC PHREG, SAS v9.4; SAS Institute, Cary, NC) was applied as a marginal model for repeated Ct values, treating Ct values as right-censored at the assay detection limit (Ct = 40). Models were stratified by target gene (N1, N2) and replicate and used robust (sandwich) variance estimation clustered by participant to account for within-subject correlation in repeated measures. Hazard ratios compare the relative hazard of an earlier event (lower Ct → higher viral RNA) for each study mouthwash vs distilled water.

Linear mixed-effects regression models (LMMs) with the least squares means and Šidák-adjusted pairwise comparisons were used to evaluate differences in Ct values among sample types (pre- and post-rinse unstimulated saliva, throat wash, and throat swab). Agreement between pre- and post-rinse values was assessed using Lin’s Concordance Correlation Coefficient (LCCC). Longitudinal changes in viral RNA from Day 1 to Day 28 were assessed using general linear models (GLM) applied to change in Ct values to RP (ΔΔCt) values for N1 and N2, with Dunnett’s adjustment for comparisons against the distilled water control group. Short-term rinse effects at baseline (pre- vs post-rinse) were also examined using GLM and multiple comparison adjustments. Mean N1 and N2 gene values were also analyzed to examine patterns. Ct values were capped at 40 cycles, corresponding to the assay detection limit [28]. Sensitivity analyses applied multiple imputations (m=10) to generate Ct values around 40. Pearson correlation coefficients were calculated to evaluate the association between Ct values and symptom status.

## Results

### Study Participants

Study participants were recruited between 01/04/2021 and 18/10/2021, from a pool of 1,422 COVID-19-positive patients. Of these, 935 met the age requirement (≥18 years) and had a COVID-19 positive test within the prior seven days, making them eligible for further research team contact for additional eligibility screening and recruitment. A total of 870 participants were excluded for various reasons, including residing too far away, current mouthwash use, plans to receive antibody treatment, lack of interest in the study, participation in another study, work conflicts, contraindicated health conditions, technological limitations (e.g., no access to a videoconference application), antiviral medication use, or failure to respond to calls or messages. Ultimately, 65 patients met the inclusion criteria, consented to participate, and were randomized. Eleven participants withdrew post-randomization (e.g., changed minds, work conflicts), but before any study procedures, leaving 54 participants who started the study. Of these, 49 (91%) completed the 28-day study, while five withdrew before Day 28 (three opted out and two lost to follow-up). Therefore, baseline (Day 1) analyses included all 54 participants who initiated the study, whereas longitudinal analyses from Day 1 to Day 28 were restricted to the 49 participants who completed follow-up. Figure 1 presents the study’s CONSORT flow diagram (Figure 1).

**Figure 1.**
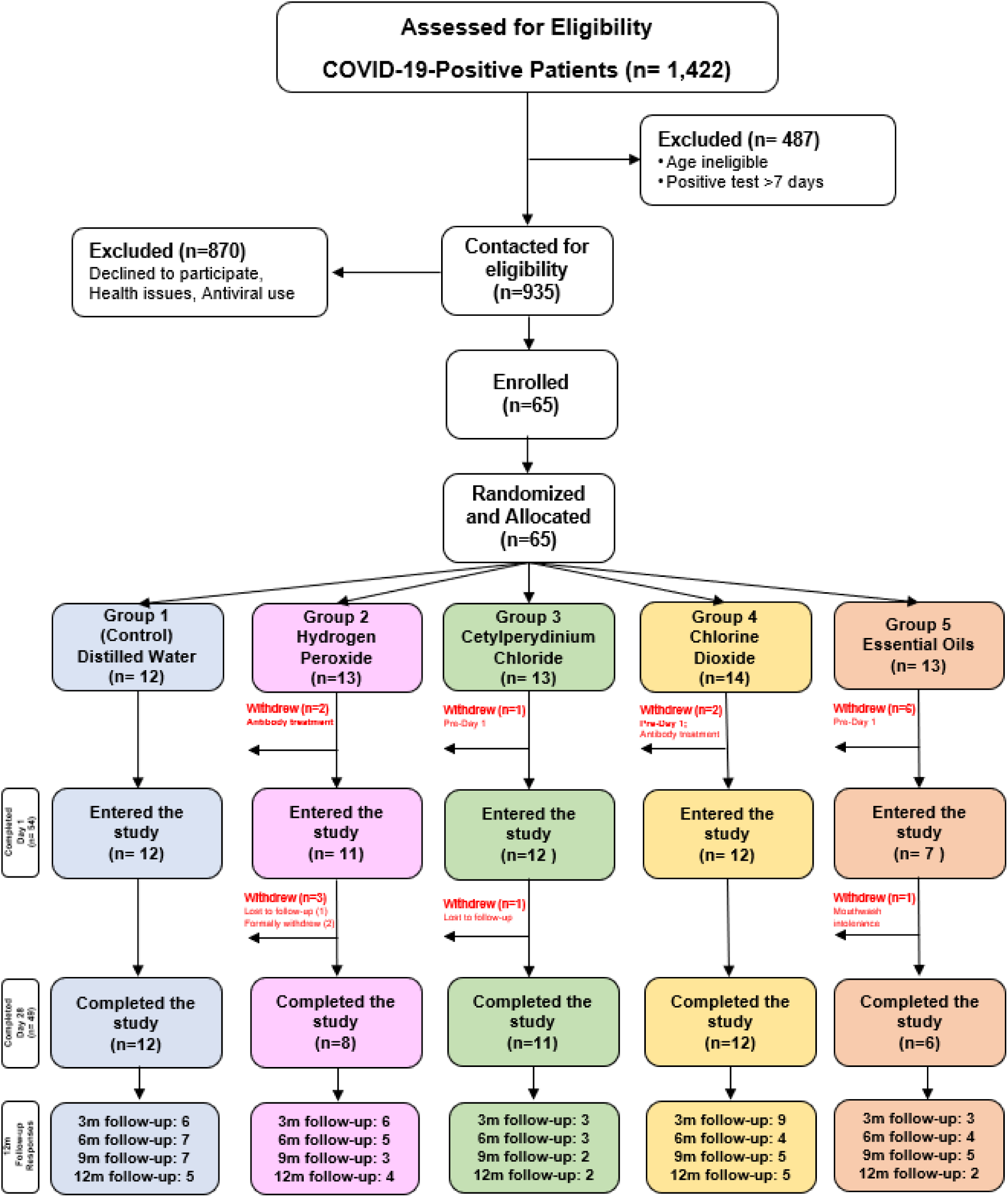
CONSORT Flow Diagram.

Retention rates were highest (92–100%) in the distilled water, chlorine dioxide, and cetylpyridinium chloride groups, while higher dropouts were observed in the hydrogen peroxide (after Day 1) and essential oils groups (before Day 1).

Table 4 presents the demographic characteristics of study participants. The mean participant age was 39.9 years. One participant (2%) was 18 years old, 48 (88%) were 19–64 years, and five (10%) were ≥65 years. The cohort was 57% female (n=31), 22% Hispanic (n=12), and 52% non-Hispanic White (n=28) (Table 4).

**Table 4.** Distribution of Study Participants across Study Groups by Demographics.

| Variable | Distilled Water | Hydrogen Peroxide | Cetylpyridinium Chloride | Chlorine Dioxide | Essential Oils | Total |
| --- | --- | --- | --- | --- | --- | --- |
| Participant Number | 12 | 11 | 12 | 12 | 7 | 54 |
| **Age (Yr)** |  |  |  |  |  |  |
| ≤ 18 | 0 | 1 | 0 | 0 | 0 | 1 |
| 19-64 | 11 | 10 | 11 | 9 | 7 | 48 |
| ≥65 | 1 | 0 | 1 | 3 | 0 | 5 |
| Mean [Standard Deviation (SD)] | 38.0 (11.5) | 31.7 (8.7) | 45.6 (11.8) | 45.3 (17.3) | 37.3 (12.5) | 39.9 (13.4) |
| **Sex** |  |  |  |  |  |  |
| Female | 8 | 8 | 5 | 6 | 4 | 31 |
| Male | 4 | 3 | 7 | 6 | 3 | 23 |
| **Race** |  |  |  |  |  |  |
| American Indian or Alaska Native | 0 | 0 | 0 | 0 | 0 | 0 |
| Asian | 4 | 1 | 2 | 2 | 0 | 9 |
| Native Hawaiian or Other Pacific Islander | 0 | 0 | 1 | 0 | 0 | 1 |
| Black or African American | 0 | 0 | 1 | 0 | 0 | 1 |
| White | 7 | 4 | 6 | 6 | 5 | 28 |
| More than one race | 1 | 0 | 0 | 3 | 0 | 4 |
| Unknown or Not Reported | 0 | 4 | 2 | 1 | 2 | 9 |
| **Ethnicity** |  |  |  |  |  |  |
| Hispanic | 0 | 4 | 2 | 3 | 3 | 12 |
| Not Hispanic | 11 | 7 | 10 | 9 | 4 | 41 |
| Unknown or Not Reported | 1 | 0 | 0 | 0 | 0 | 1 |

No participant was hospitalized due to COVID-19 during the study. Three participants, who were healthcare providers, were required to leave home for work. Only 17 participants (35%) responded to 3 or more quarterly follow-up questionnaires.

### Primary Outcome Measure: Ct Value Changes Over Time

Ct values were measured for Target N1 and N2 genes in study samples (i.e., swab, unstimulated saliva, and throat wash) at baseline (Day 1) and on Days 7 and 28, comparing pre- and post-rinse samples for each study mouthwash. Baseline (Day 1) comparisons included all 54 participants who initiated the study. Longitudinal analyses from Day 1 to Day 28 were restricted to participants who completed follow-up (n = 49). Descriptive baseline summaries include all enrolled participants (n = 54).

#### Comparison of Ct Value in Baseline (Day 1) Samples

Ct values from five samples (i.e., throat swab, and pre- and post-rinse unstimulated saliva and throat wash) on Day 1 were compared using a linear mixed-effects model (LMM) with Šidák-adjusted pairwise comparisons. Results showed that pre-swab samples had significantly lower Ct values than pre-unstimulated saliva and throat wash samples (P < 0.001), suggesting that swab collection captures higher viral RNA levels than the other two sampling methods. Post-rinse unstimulated saliva and throat wash samples had significantly higher Ct values (lower viral load) than pre-swab samples (P < 0.001), likely due to dilution effects from rinsing. However, no significant differences were observed between pre- and post-rinse samples within the same collection method (i.e., unstimulated saliva or throat wash), over the short (5-minute) post-rinse interval. This indicates that rinsing did not immediately result in statistically significant viral RNA level reductions (Table 5).

**Table 5.** Comparison of Ct Value of Day 1 Samples (N=54)

| Samples | Ct Value – Mean (SD) |  |
| --- | --- | --- |
|  | N1 | N2 |
| Pre-Swab vs. Pre-Unstimulated Saliva | 27 (0.5) vs. 31 (0.5) □ | 27 (0.5) vs. 31 (0.5) □ |
| Pre-Swab vs. Pre-Throat Wash | 27 (0.5) vs. 30 (0.5) □ | 27 (0.5) vs. 30 (0.5) □ |
| Pre-Swab vs. Post-Unstimulated Saliva | 27 (0.5) vs. 32 (0.5) □ | 27 (0.5) vs. 32 (0.5) □ |
| Pre-Swab vs. Post-Throat Wash | 27 (0.5) vs. 30 (0.5) □ | 27 (0.5) vs. 30 (0.5) □ |
| Pre-Unstimulated Saliva vs. Pre-Throat Wash | 31 (0.5) vs. 30 (0.5) | 31 (0.5) vs. 30 (0.5) |
| Pre-Unstimulated Saliva vs. Post-Unstimulated Saliva | 31 (0.5) vs. 32 (0.5) | 31 (0.5) vs. 32 (0.5) |
| Pre-Unstimulated Saliva vs. Post-Throat Wash | 31 (0.5) vs. 30 (0.5) | 31 (0.5) vs. 30 (0.5) |
| Pre-Throat Wash vs. Post-Throat Wash | 30 (0.5) vs. 30 (0.5) | 30 (0.5) vs. 30 (0.5) |
| a: P < 0.001; Ct values were rounded to closest integer; SD=standard deviation |  |  |

#### Agreement Between Baseline Sample Types (LCCC Analysis)

The LCCC was used to evaluate the concordance of Ct values among the five baseline sample types (Table 3). As Table 6 shows, when comparing pre-rinse unstimulated saliva to pre-swab samples, the agreement was moderate. In contrast, pre-rinse throat wash exhibited somewhat stronger agreement with pre-swab samples. Post-rinse throat wash samples continued to show moderate agreement with pre-swab Ct values, while post-rinse unstimulated saliva showed weaker agreement with pre-swab samples. These results suggest that throat wash may provide a more reliable measurement of SARS-CoV-2 viral load compared to unstimulated saliva when swab collection is not feasible.

**Table 6.** Lin’s Concordance Correlation Coefficient (LCCC) for Pre- and Post-Rinse Ct Values by Target (N=49)

|  |  |  |  |  |
| --- | --- | --- | --- | --- |
| **Comparison** | **Target** | **LCCC** | **95% CL** | **95% UL** |
| Pre-Saliva<sup>A</sup> vs Pre-Swab<sup>B</sup> | N1 | 0.56 | 0.39 | 0.72 |
|  | N2 | 0.59 | 0.43 | 0.75 |
|  | RP | 0.06 | -0.05 | 0.16 |
| Pre-TW<sup>C</sup> vs Pre-Swab | N1 | 0.69 | 0.56 | 0.82 |
|  | N2 | 0.70 | 0.57 | 0.82 |
|  | RP | 0.07 | -0.05 | 0.20 |
| Post-Saliva<sup>D</sup> vs Pre-Saliva | N1 | 0.83 | 0.74 | 0.91 |
|  | N2 | 0.82 | 0.73 | 0.91 |
|  | RP | 0.47 | 0.28 | 0.66 |
| Post-TW<sup>E</sup> vs Pre-TW | N1 | 0.90 | 0.85 | 0.95 |
|  | N2 | 0.90 | 0.85 | 0.95 |
|  | RP | 0.57 | 0.39 | 0.76 |
| Post-Saliva vs Pre-Swab | N1 | 0.50 | 0.33 | 0.67 |
|  | N2 | 0.51 | 0.34 | 0.68 |
|  | RP | 0.06 | -0.09 | 0.21 |
| Post-TW vs Pre-Swab | N1 | 0.72 | 0.60 | 0.84 |
|  | N2 | 0.74 | 0.63 | 0.85 |
|  | RP | 0.01 | -0.12 | 0.14 |

A: Pre-rinse unstimulated saliva; B: Pre-rinse swab; C: Pre-rinse throat wash; D: Post-rinse unstimulated saliva E: Post-rinse throat wash

#### Effect of Study Mouthwashes on Ct Values Over Time

A Cox proportional hazards regression model was used to evaluate the effect of different mouthwashes on SARS-CoV-2 viral load over time, applying a marginal model for repeated Ct values and stratifying by target gene (N1, N2). This modeling framework enabled estimation of relative viral load dynamics while accounting for censoring at the assay detection limit and within-subject correlation using robust (sandwich) variance estimation clustered by participant. No significant differences in Ct values were observed among study arms during the study period (P > 0.05). Hazard ratios (HRs) compared with distilled water ranged from 0.82 to 0.86; however, these differences were not statistically significant (P > 0.05), indicating no evidence of a sustained treatment effect over time.

An exception was observed on Day 7, when cetylpyridinium chloride demonstrated a statistically significant association (HR = 0.36, 95% CI 0.21–0.62; P = 0.001), consistent with a transient increase in Ct values (i.e., reduced viral RNA). No significant differences were observed on Day 28.

Figure 2 shows mean Ct values on Day 1 across study groups for five sample types: pre-rinse swab, pre- and post-rinse unstimulated saliva, and pre- and post-rinse throat wash. Across groups, pre-rinse samples generally exhibited lower Ct values than post-rinse samples, consistent with a short-term increase in Ct values following rinsing (reflecting reduced detectable viral RNA after rinsing). However, no statistically significant differences in Ct values were observed among study groups at baseline (P > 0.05). Similarly, no significant differences were observed in pre-rinse throat wash samples on Day 1 between study groups and the control group (P = 0.75).

**Figure 2.**
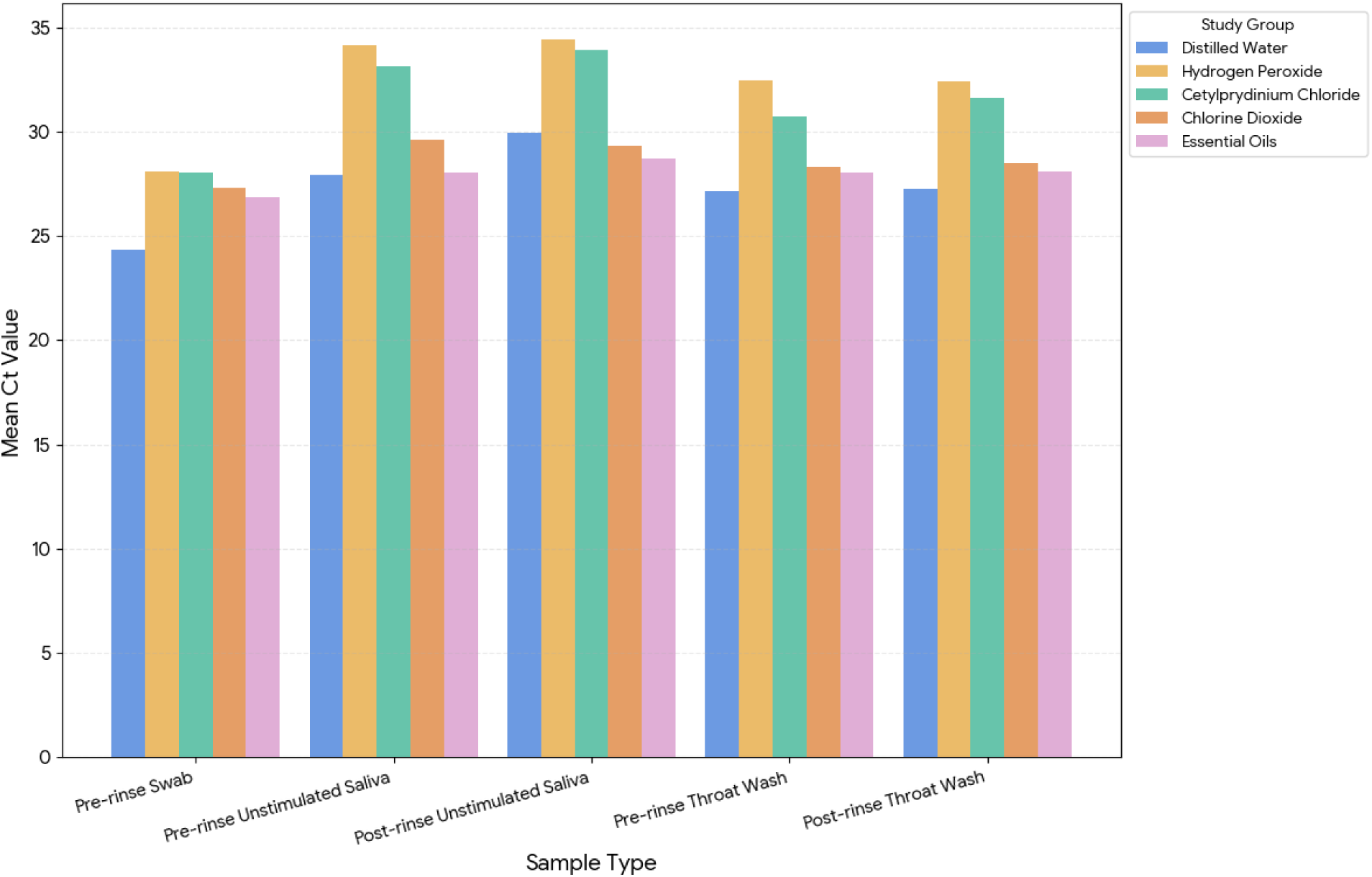
Comparison of Ct Values of Day 1 Samples among Study Groups (N=54)

As shown in Figure 3, RP Ct values remained relatively stable over time, whereas viral targets (N1, N2, and the mean of N1 and N2) showed progressive increases in Ct values (>32 on Day 7 and >35 on Day 28), consistent with natural viral clearance. Although cetylpyridinium chloride exhibited higher Ct values on Day 7 compared with other groups, no significant differences were observed by Day 28, suggesting that observed changes likely reflect the expected course of infection rather than a sustained mouthwash-specific effect.

**Figure 3.**
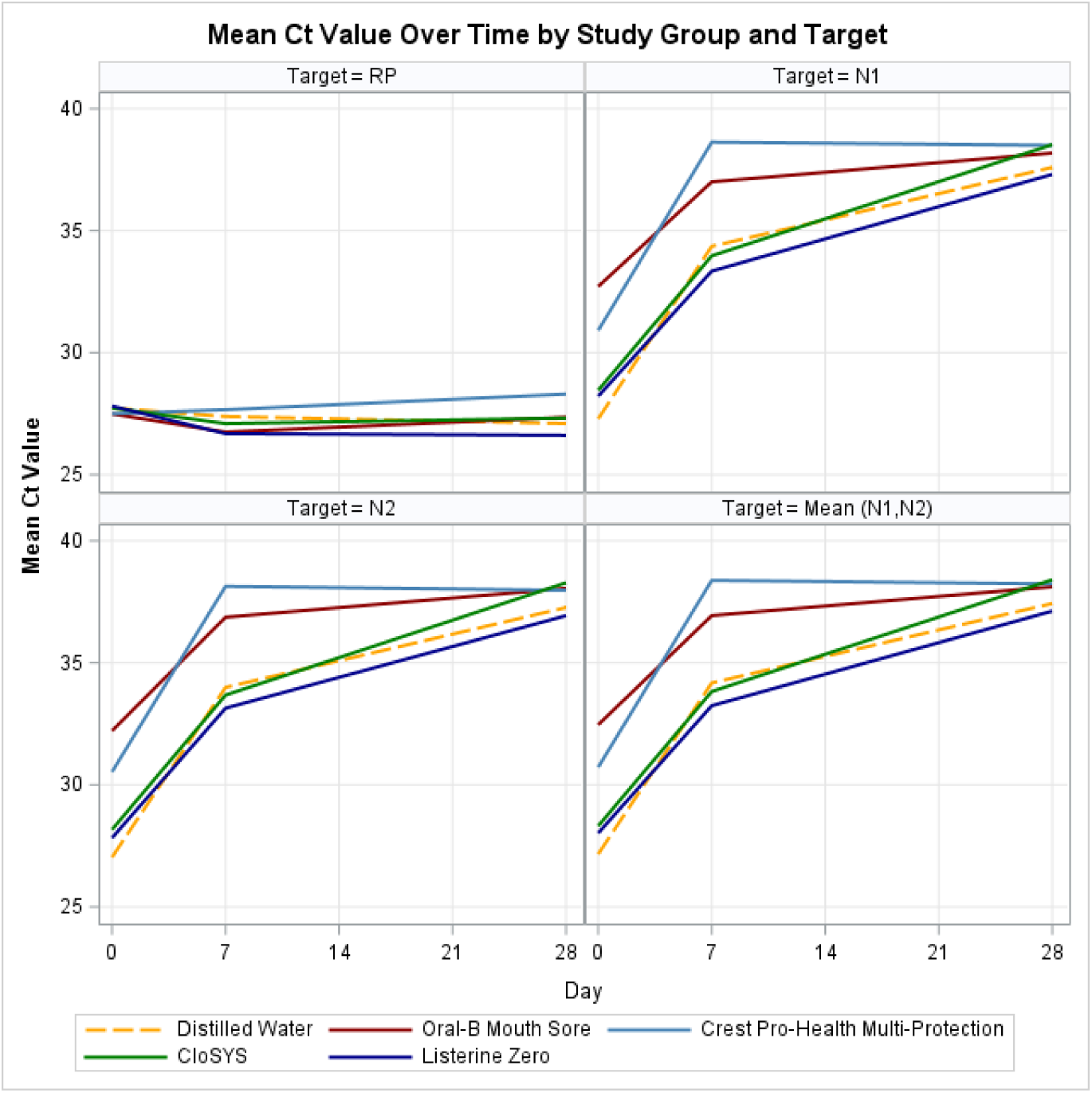
Mean Ct Value Over Time by Study Group for Throat Wash Samples.

#### Comparison of Pre-Rinse Throat Wash Samples Over Time

Pre-rinse throat wash samples were collected on Days 1, 7, and 28, while other sample types were collected only on Day 1. Table 7 summarizes the mean change in Ct values (Targets N1 and N2) for pre-rinse throat wash samples across study groups from baseline to Day 28. Chlorine dioxide and essential oils showed somewhat greater increases in Ct values compared to the distilled water, suggesting possible reductions in viral RNA. However, no differences were statistically significant when compared to the control group (P > 0.05). Notably, the control group also showed a substantial increase in Ct values over time, indicating a natural decline in viral load as infections resolved. These findings suggest that observed changes in Ct values likely reflect the expected course of SARS-CoV-2 infection rather than the treatment effect of the study mouthwashes.

**Table 7.** Mean (SD) Change in Cycle Threshold (Ct) Values (Targets N1 and N2) of Pre-Rinse Throat Wash Samples from Day 1 (Baseline) to Day 28 Compared with Distilled Water (N=49)

| Mouthwash<br>Variable | Distilled<br>Water | Hydrogen<br>Peroxide | Cetylpyridinium<br>Chloride | Chlorine<br>Dioxide | Essential<br>Oils | Total |
| --- | --- | --- | --- | --- | --- | --- |
| Number of<br>participants | 12 | 8 | 11 | 12 | 6 | 49 |
| Change in Ct | 10.3 (5.1) | 6.1 (6.0) | 7.3 (5.6) | 10.1 (7.9) | 10.6 (5.4) | 8.9 (6.2) |
| **(target N1)**<br>(undetermined Ct=40) |  | -4.2 (-11.4, 3.1)<br>P=0.41 | -3.1 (-9.6, 3.6)<br>P=0.62 | -0.2 (-6.7, 6.2)<br>P=1.0 | 0.3 (-7.6, 8.2)<br>P=1.0 |  |
| **Change in Ct**<br>**(target N2)**<br>(undetermined Ct=40) | 10.2 (5.4) | 6.6 (6.0)<br>-3.7 (-11.1, 3.7)<br>P=0.55 | 7.1 (6.1)<br>-3.1 (-9.8, 3.7)<br>P=0.62 | 10.1 (7.9)<br>-0.1 (-6.7, 6.5)<br>P=1.0 | 10.3 (5.2)<br>0.1 (-8.0, 8.2)<br>P=1.0 | 8.9 (6.3) |
| **Change in Ct**<br>**(target N1 and N2 combined)**<br>(undetermined Ct=40) | 10.3 (5.2) | 6.3 (6.0)<br>-3.9 (-11.2, 3.4)<br>P=0.48 | 7.2 (5.9)<br>-3.1 (-9.7, 3.6)<br>P=0.62 | 10.1 (7.9)<br>-0.1 (-6.7, 6.5)<br>P=1.0 | 10.5 (5.3)<br>0.2 (-7.8, 8.2)<br>P=1.0 | 8.9 (6.2) |
| The first value in each cell represents the mean change in Ct (± SD) from baseline (Day 1) to Day 28. The second value represents the adjusted difference in mean change compared with the distilled water control group (95% CI), with corresponding Dunnett-adjusted P values. Ct values were capped at 40 cycles, representing the assay detection threshold. | | | | | | |

To further evaluate changes in viral RNA levels, a multiple comparison analysis was performed using ΔΔCt values, which measure changes in Ct values over time. ΔΔCt presents differences in Ct values between the target gene (N1 or N2) and RP from baseline to Day 28. Higher ΔΔCt values correspond to greater reductions in viral RNA, while a lower or negative value suggests little or no change. The overall model was not significant for either N1 vs RP (p = 0.2197) and N2 vs RP (p = 0.2667), indicating no strong evidence that any of the rinses reduced viral RNA compared to the control group. Dunnett’s adjusted pairwise comparisons confirmed that none of the interventions significantly differed from the control group (Figure 4). Least Squares Means analysis showed that the mean ΔΔCt for the control group was 10.94 (N1 vs RP) and 10.87 (N2 vs RP), with chlorine dioxide and essential oils having similar values and no statistically significant differences (P > 0.05) (Appendix C).

**Figure 4.**
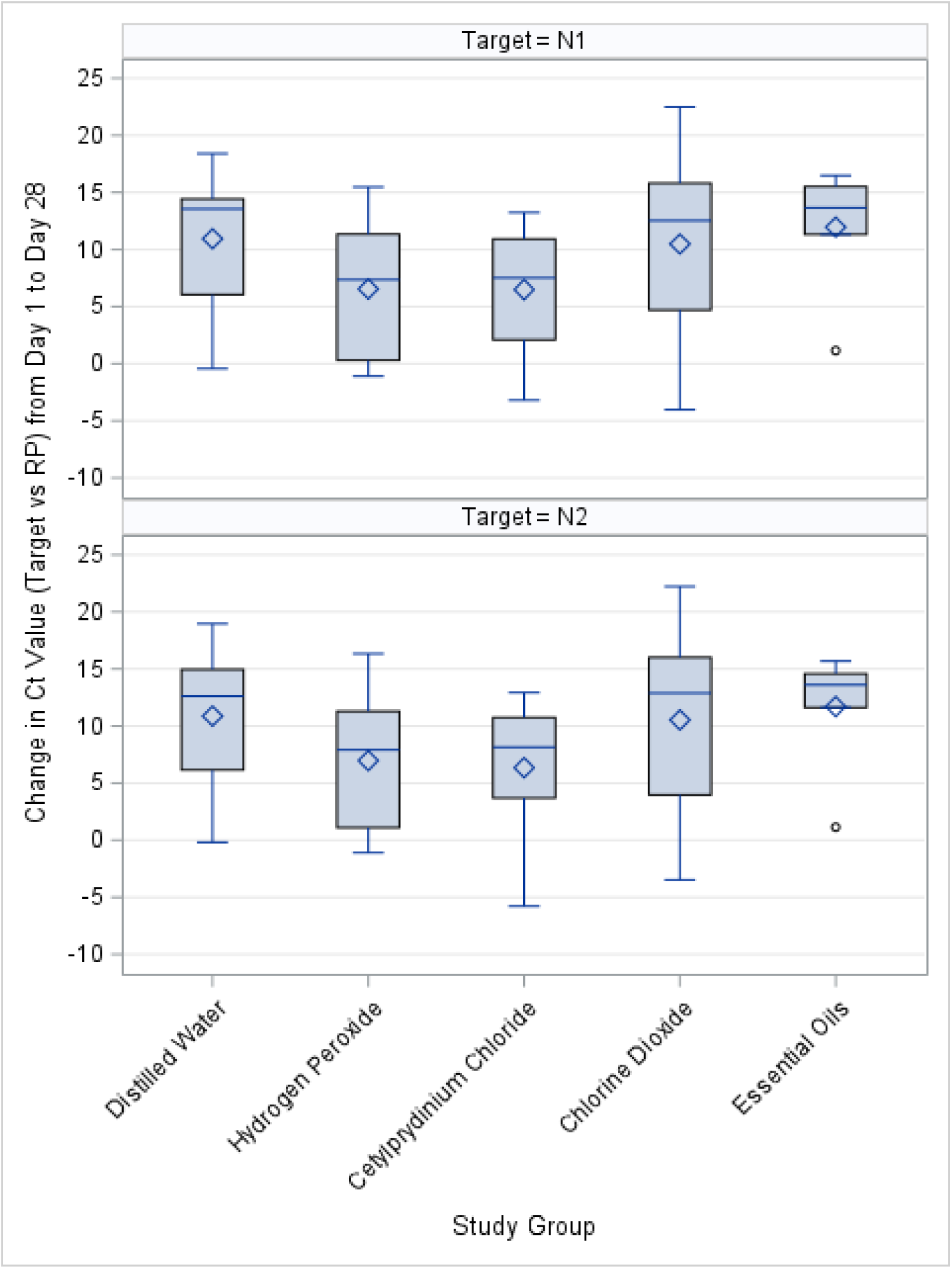
Change in Ct Values from Day 1 to Day 28 by Study Group. Box-and-whisker plots show the change in Ct values (target Ct normalized to RP) between Day 1 and Day 28 for the N1 target (top panel) and N2 target (bottom panel). Boxes represent the interquartile range with the median indicated by the horizontal line; diamonds denote the mean; whiskers represent the range, and circles indicate outliers. Positive values indicate an increase in Ct (corresponding to lower detectable viral RNA over time).

### Secondary Outcome Measures

The secondary outcome measures included self-reported COVID-19 clinical symptoms (baseline to three months) and any healthcare utilization/hospitalization (baseline to four weeks).

*<u>a. COVID-19 Related Symptoms.</u>* Of 54 participants, 46 (85%) were symptomatic at baseline (Day 1), with variation across study arms and eight (15%) were asymptomatic (Figure 5).

**Figure 5.**
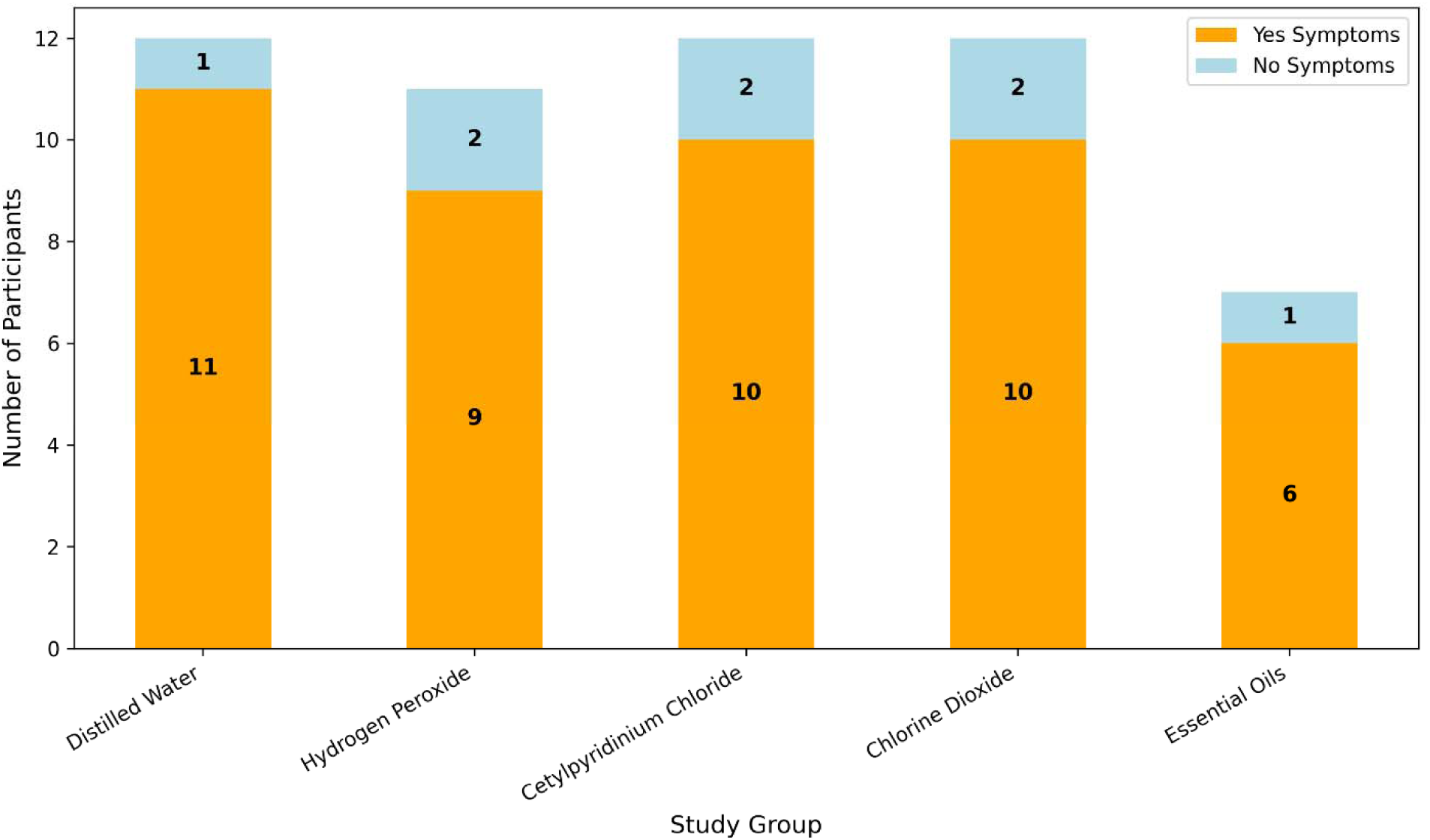
Frequency of Symptomatic and Asymptomatic Participants by Study Group on Day 1 (Baseline)

The most common symptoms were nasal congestion (n=34, 63%), cough (n=26, 48%), lost taste or smell (n=19, 35%), headache (n=23, 43%), and fatigue (n=19, 35%). (Figure 6).

**Figure 6.**
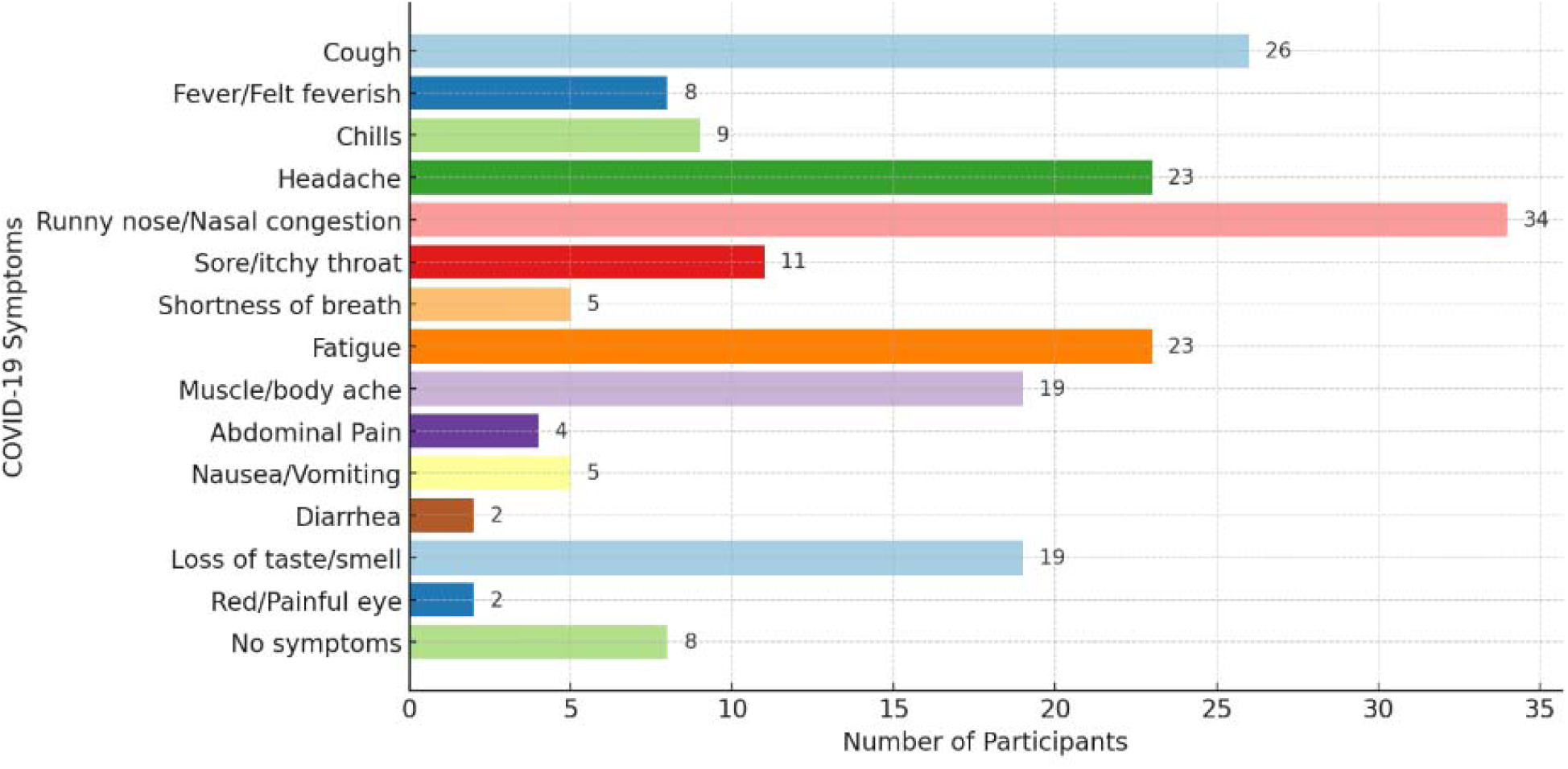
COVID-19 Related Symptoms Among Participants on Day 1 (Baseline)

The Pearson correlation between “still being symptomatic” and pre-rinse Ct values showed negative correlations, meaning that lower Ct values were associated with more symptoms. The strongest correlation was seen with pre-rinse swab Ct values (−0.34, P < 0.012), suggesting that participants with lower Ct values were more likely to be symptomatic. Unstimulated saliva and throat wash Ct values had weaker correlations with symptoms and were not statistically significant (P > 0.05).

*<u>b. Healthcare Utilization/Hospitalization</u>.* No participants experienced progression or clinically meaningful changes in their health conditions requiring healthcare utilization or hospitalization during the study period. Consequently, the planned moderator analyses evaluating the impact of healthcare utilization or hospitalization on the primary outcome were not conducted.

### Additional Outcomes

#### Tobacco Product Use

Baseline tobacco-use data were collected using items from the Population Assessment of Tobacco and Health (PATH) questionnaire. Exploratory descriptive analyses examined whether tobacco use, marijuana smoking, or vaping were associated with SARS-CoV-2 viral load at baseline or during follow-up (Appendix D). Across all tobacco- and nicotine-related variables—including cigarette use in the past 12 months or 30 days, e-cigarette use, and cigar/shisha/hookah use—Pearson correlation coefficients with Ct values at each time point were small in magnitude and not statistically significant (all P > 0.05). Similar patterns were observed across all three RT-qPCR targets (N1, N2, and RP). Because very few participants reported tobacco, marijuana, or vaping behaviors, no inferential subgroup analyses or moderator analyses were conducted (Appendix D)

### Additional Self-Reported Characteristics

#### Symptoms or Issues with Assigned Mouthwash

Participants reported symptoms or issues through daily questionnaires. Across all study arms, 17 participants (31%) experienced problems with their assigned mouthwash, while 37 (69%) reported no issues. The most frequently reported issue was difficulty adhering to mouthwash usage guidelines (11 mentions), followed by challenges with gargling (6 mentions) and oral blisters (5 mentions). Additional complaints included burning sensations, confusion about instructions, and discomfort with aftertaste (Appendix E). Some issues were attributable to participant error, whereas others were related to the mouthwash formulation. The percentage of participants reporting issues varied across groups, ranging from 43% in the essential oils group (3 of 7) to 25% in both the chlorine dioxide and cetylpyridinium chloride groups (3 of 12 each). Notably, six participants in the cetylpyridinium chloride group found its foamy texture unpleasant, making it difficult to retain in the mouth for the manufacturer-recommended duration. Two participants disliked the mint flavor of the hydrogen peroxide mouthwash. It is possible that participants who discontinued the study may not have fully reported complaints about their assigned mouthwash.

#### Pre-Existing Health Conditions

At baseline, among the 54 study participants, 38 (∼70%) reported no pre-existing health conditions, while eight participants reported unspecified conditions. Of those with identified conditions, five participants reported cardiovascular disease, three reported pulmonary disease, and three reported diabetes mellitus. Additional conditions reported included atrial fibrillation, osteoporosis, arthritis, anemia, adenomyosis, epilepsy, Post-Traumatic Stress Disorder (PTSD), migraines, fibromyalgia, anxiety, atopic dermatitis, and mild adrenal insufficiency. Participants with existing conditions continued their prescribed medications throughout the study. No changes in health conditions were reported during the study period. Although a moderator analysis was planned, it was not conducted due to the small number of participants with documented conditions.

#### Vaccination Status

Of the 54 participants, 45 (83%) had received both doses of a COVID-19 vaccine, three (6%) had received only the first dose, and six (11%) were unvaccinated. Of the 49 participants who completed the study from Day 1 to Day 28, 32 (65%) received both doses of the Pfizer COVID-19 vaccine, 10 (20%) received both doses of Moderna, two (4%) received one dose of Johnson & Johnson, and five (10%) were unvaccinated. None of the participants received more than one vaccine type.

#### Oral Hygiene Practices

At baseline, participants were asked about their oral hygiene practices. More than half (52%) reported brushing their teeth twice daily, 37% flossed once daily, 52% did not use any mouthwash, and 65% did not gargle. Regarding tooth loss, 19 participants (35%) had partial tooth loss (1–6 teeth), one participant (2%) was edentulous, and 34 participants (63%) had no tooth loss (Appendix F).

## Discussion

This randomized double-masked controlled trial evaluated whether four OTC antiseptic mouthwashes reduced SARS-CoV-2 viral load compared with distilled water, using Ct values from RT-qPCR across multiple sample types. Ct values increased over time across all groups—including the control—indicating natural viral clearance, consistent with findings reported by Badu et al [29]. We did not observe statistically significant differences among study arms, aside from a transient reduction in viral load on Day 7 in the cetylpyridinium chloride group, which had slightly higher Ct values at baseline (Day 1) than the distilled water control. Variations in Ct values across sample types underscore the importance of sampling method in SARS-CoV-2 detection.

Beyond the clinical findings, this study presents a longitudinal analytical framework for evaluating RT-qPCR Ct values in outpatient viral infections. By modeling Ct values as censored time-to-event outcomes, we incorporated the assay detection limit and repeated within-participant measurements into a unified analytic approach. Such frameworks may be informative for future clinical trials evaluating antiviral interventions using RT-qPCR–based endpoints, particularly when detection thresholds complicate conventional linear analyses. The following sections integrate these findings with existing literature and discuss their clinical relevance.

### Effect of Study Mouthwashes on SARS-CoV-2 Load Over Time

Statistical analyses showed no significant differences between any mouthwash and distilled water at any time point. The temporary reduction in viral load observed with cetylpyridinium chloride on Day 7 was not sustained at Day 28. This aligns with prior studies demonstrating inconsistent or minimal in vivo effects of cetylpyridinium chloride despite promising in vitro virucidal activity. In a randomized, placebo-controlled trial of adults with asymptomatic or mild COVID-19, a single rinse with 0.05% cetylpyridinium chloride or 0.01% aqueous chlorine dioxide did not significantly reduce salivary SARS-CoV-2 levels at any time point up to 24 hours compared with a water rinse, suggesting that under these conditions neither agent effectively lowers oral viral load [30].

Ferrer et al. conducted a multi-center, double-masked, randomized trial underscoring the limited in vivo efficacy of common antiseptic rinses against SARS-CoV-2; despite robust in vitro virucidal activity, neither povidone-iodine, hydrogen peroxide, 0.07% cetylpyridinium chloride nor chlorhexidine produced a meaningful reduction in salivary viral RNA up to two hours post rinse-, with placebo and treatment arms exhibiting similar transient declines [31]. Similarly, another randomized controlled trial reported stable

SARS-CoV-2 RNA levels up to 60 minutes post-use of 0.05% cetylpyridinium chloride mouthwash [30]. Systematic reviews also suggest that 0.075% cetylpyridinium chloride mouthwashes may not effectively reduce viral load, likely because cetylpyridinium chloride disrupts the lipid membrane of the viral envelope without affecting viral RNA integrity [32]. Tarragó-Gil et al. found no significant reduction in viral RNA 120 minutes after rinsing with 0.07% cetylpyridinium chloride; however, they reported increased viral lysis, suggesting a potential impact on infectivity [33].

Alemany et al. conducted a multicenter randomized clinical trial evaluating a single rinse with 0.07% cetylpyridinium chloride mouthwash in patients with mild to moderate COVID-19.34 While RT-qPCR did not show significant reductions in viral RNA, ELISA detected increased salivary nucleocapsid (N) protein concentrations in the cetylpyridinium chloride group compared with placebo, suggesting that cetylpyridinium chloride may compromise viral structural integrity and reduce infectivity even when viral RNA remains detectable [34].

Bezinelli et al. found a formulation combining 0.075% cetylpyridinium chloride with zinc lactate significantly reduced viral load for up to 60 minutes, highlighting the importance of formulation and additives in enhancing efficacy [35]. Onozuka et al. found that low concentrations of cetylpyridinium chloride (0.001–0.005%) suppressed SARS-CoV-2 infectivity in saliva without disrupting the viral envelope, suggesting that cetylpyridinium chloride may act by denaturing viral proteins rather than solely relying on lipid membrane degradation. These findings highlight the need for further research on cetylpyridinium chloride concentrations, formulations, and their effects on viral components to determine their potential role in mitigating SARS-CoV-2 transmission. Additionally, natural oral clearance from saliva swallowing may contribute to an eventual resurgence of active viral particles in the oral cavity [30].

Our study results align with Al Zahrani et al. regarding the hydrogen peroxide effectiveness on SARS-CoV-2 load, reporting that viral load was reduced at 5-, 30-, and 60-minutes relative to no rinse, but was not significantly different from distilled water at baseline [36]. The slight decrease in viral load observed in our distilled water group suggests mechanical clearance, a finding also reported by Al Zahrani et al [36]. Perussolo et al. evaluated the efficacy of 0.2% chlorhexidine, 1.5% hydrogen peroxide, and 0.07% cetylpyridinium chloride mouthwashes in reducing SARS-CoV-2 viral load in hospitalized COVID-19 patients. Saliva samples were collected after 30 seconds of mouthwash rinsing at baseline and up to 3 hours post-rinsing. While all groups, including the control, showed viral load reductions at 2 and 3 hours, no significant differences were observed between mouthwashes and the control groups [37].

### Sample Type and SARS-CoV-2 Viral Load

At the onset of the COVID-19 pandemic, researchers explored various sampling methods to detect SARS-CoV-2 reliably. Studies have since compared saliva (both stimulated and unstimulated), nasopharyngeal and oropharyngeal swabs, bronchoalveolar-lavage fluid, and throat wash (gargle lavage) [37–39]. Zhu et al. highlighted saliva’s role in monitoring viral load and transmission [40]. Sakanashi et al. found that saliva had higher sensitivity than nasopharyngeal swabs in COVID-19 patients [41]. Wyllie et al. reported that self-collected saliva showed greater sensitivity and lower variability than nasopharyngeal swabs [42].

We compared five sampling methods at baseline to evaluate the difference between various methods in reliably collecting viral samples. Our findings showed that pre-rinse throat swab samples had the highest viral load. There was also strong agreement between pre-rinse throat swab and pre- and post-rinse throat wash samples, suggesting that throat-based sampling may provide a more reliable alternative to saliva. Additionally, the analysis of pre-rinse throat wash samples collected at baseline (Day 1), Day 7, and Day 28 showed greater changes in Ct value in the chlorine dioxide and essential oils mouthwash groups. However, these changes were similar to those in the control group, indicating no significant effect on viral load.

These results align with prior research indicating that SARS-CoV-2 viral loads tend to be higher in samples obtained from deeper respiratory tract locations (29). Although our study did not include lower respiratory tract (LRT) samples, instructing participants to swab the posterior pharyngeal wall ensured that throat swab samples were collected from a deeper URT site than unstimulated saliva in the oral cavity. Additionally, throat wash sampling may capture viral particles from LRT secretions via mucociliary transport, further supporting lower Ct values in throat-based methods [29].

Onozuka et al. collected unstimulated saliva samples by swabbing participants’ mouths at multiple time points from baseline to 24 hours. Their study found that neither cetylpyridinium chloride nor aqueous chlorine dioxide mouthwash significantly reduced viral load compared to the placebo (purified water) [30]. These findings highlight the need to evaluate multiple sampling and detection methods to determine the most reliable approach for SARS-CoV-2 testing.

### Mouthwash Use and Clinical Symptoms

Participants with more symptoms had lower Ct values, consistent with the literature linking higher viral load to greater symptom burden [43]. The most frequently reported symptoms - runny nose/nasal congestion, cough, headache, loss of taste/smell, and fatigue - align with the well-documented COVID-19 symptomatology. Symptoms resolved over time, reflecting the natural disease course.

A few studies have shown the mouthwash effect on COVID-19 symptom prevention or alleviation. Karami et al. investigated the effect of different mouthwashes, including 0.2% chlorhexidine and 7.5% sodium bicarbonate, and found that chlorhexidine significantly reduced symptom incidence among healthcare workers [44]. While our study did not assess the same mouthwashes and symptom onset as an outcome measure, it does indicate symptom resolution over time, which may be attributed to mouthwash use or the natural course of disease recovery.

Poleti et al. evaluated the effects of an antimicrobial phthalocyanine derivative (APD) in mouthwash and dentifrice on COVID-19 symptoms in a randomized, triple-masked clinical trial [45]. Participants followed an oral hygiene regimen for seven days. While both groups showed symptom reduction, the APD group had a greater decrease in cough, fatigue, shortness of breath, loss of smell, altered taste, sore throat, and other symptoms. On days three and seven, the control group had a higher symptom prevalence. The findings suggest that APD-containing oral hygiene products may help reduce COVID-19-related symptoms [45]. Future research should conduct clinical trials that assess both virological and symptomatic outcomes in patients using antiviral mouthwashes.

### Implications for Clinical Guidelines on Mouthwash Use

Organizations such as ADA and CDC recommended pre-procedural rinses early in the pandemic despite limited clinical evidence [19,20]. However, our findings do not support a meaningful reduction in SARS-CoV-2 viral load with hydrogen peroxide or any other tested mouthwash.

Additionally, no tested mouthwash, including distilled water, exhibited significant antiviral properties. These findings contrast with prior in vitro testing of the same mouthwash formulations conducted by researchers in Germany, which demonstrated significant antiviral activity—particularly for essential oils and other compounds—within 30 seconds under controlled laboratory conditions. The discrepancy underscores the difference between in vitro virucidal activity and clinical effectiveness in vivo [21]. Salivary components, such as glycoproteins, are likely to interfere with the mouthwashes’ antimicrobial activity, neutralize or interfere with the active ingredients of mouthwashes, diminishing their efficacy, and undergo a 1:4 dilution due to the presence of saliva, potentially reducing the mouthwash effect [31]. Studies on mouthwashes used in our study have shown their binding to other microorganisms in the oral cavity; thus, a lower product availability *in vivo* is expected [46]. Schwarz et al. (2021) showed that oral biofilms significantly reduce the effectiveness of commonly used antiseptics due to limited penetration and microbial adaptation [47]. On the other hand, SARS-CoV-2 is shed not only in saliva but also from deeper respiratory sites, suggesting that mouthwashes may not reach all viral reservoirs, and continuous viral shedding could quickly replenish viral loads [29]. Moreover, the short contact time of mouthwashes *in vivo* and dilution by saliva may reduce their effectiveness compared to controlled laboratory conditions [29]. Host immune factors, including antibodies and antimicrobial peptides, could also influence the virus’s persistence [48]. Furthermore, *in vitro* experimental conditions do not fully replicate real-world variables like temperature fluctuations, pH changes, and biological variability among patients [48]. These interactions highlight the complexity of the oral environment and suggest that salivary constituents can modulate the effectiveness of mouthwash formulations.

### Participant Health Profiles

Study participants were diverse with varying demographic and health characteristics. The symptom burden observed in this study was consistent with reports from other outpatient COVID-19 cohorts, reinforcing the relevance of our findings. An important finding was the relationship between symptom persistence and viral load. Lower Ct values in pre-swab samples were significantly correlated with prolonged symptoms, while saliva and throat wash Ct values were weaker predictors of symptom duration. These findings suggest that pre-swab sampling may provide more consistent viral detection. Previous research has demonstrated that higher viral loads are associated with increased symptom severity and prolonged viral shedding [29].

Individual differences in host response may also have contributed to variability in observed Ct patterns [48]. While adherence to the mouthwash regimen was self-reported and supported by daily reminders, some participants occasionally forgot to use the product as instructed. Such variations in adherence, along with differences in baseline viral load and immune response, may have contributed to the variability observed in Ct value changes over time.

#### Impact of Tobacco, Marijuana, and Vape Use

No meaningful associations were observed between tobacco exposure and viral load, echoing findings from other studies showing limited direct effects on oral SARS-CoV-2 viral dynamics [49]. Larger studies are needed to clarify dose-dependent or long-term effects.

### Oral Hygiene Practices

Although not a focus of this trial, we collected oral hygiene data rarely captured in prior COVID-19 studies. Gregorczyk-Maga et al. assessed toothbrushing practice in hospitalized COVID-19 patients, showing the positive impact of toothbrushing on reducing different bacteria in the oral cavity [50]. Future research should explore whether routine hygiene behaviors influence viral shedding.

### Mouthwash Use Compliance and Challenges

At baseline (Day 1), participants received guidance on proper mouthwash-use techniques and daily reminders to support adherence. Nevertheless, compliance varied, with several participants reporting difficulty maintaining consistent use due to factors such as forgetting doses despite reminders, irregular work schedules, and tolerability issues, including unfavorable flavor, foamy texture, burning sensations, and blistering. Differences in retention between study groups may reflect these tolerability challenges, particularly in groups where participants reported more discomfort or dislike of the assigned mouthwash (hydrogen peroxide and essential oils groups). Similar findings have been reported by Natto et al., where adherence to chlorhexidine mouthwash was affected by its bitter taste [51]. These observations highlight the importance of considering product acceptability, user preferences, and ongoing bidirectional communication to enhance adherence and retention in clinical trials.

Despite the insightful findings, this study had several limitations. The study protocol was originally developed in 2020 at the onset of the COVID-19 pandemic, but its execution faced multiple challenges. Initially, the research team planned to include povidone-iodine mouthwash as one of the study arms; however, because it was not FDA-approved for mouthwash use in the US, the UCSF IRB did not approve its inclusion as a study arm. This resulted in delays associated with revising and resubmitting the study application. Furthermore, the study was initially designed for in-person sample collection, but worsening pandemic conditions and institutional mandatory remote work for research whenever possible required a transition to online sessions. Participants were guided virtually through the sample collection process, which led to additional delays and logistical challenges. Recruitment efforts were also affected by introducing COVID-19 vaccines and monoclonal antibody treatments, which rendered many potential participants ineligible or led to their withdrawal after recruitment. Despite regular reminders, some participants occasionally forgot to use the mouthwash as directed, impacting adherence to the protocol. The small sample size (n = 54) limited the study’s statistical power, reducing the ability to detect subtle differences between groups. A larger sample would have allowed for a more comprehensive evaluation of mouthwash efficacy.

In addition, individual variability in viral clearance—driven by differences in immune response, symptom severity, and stage of infection—may have contributed to heterogeneity in Ct values. In addition, the 28-day follow-up may not have been sufficient to evaluate longer-term effects of repeated mouthwash use. Finally, reliance on self-reported adherence introduces the potential for recall bias and variability in compliance.

### Future Directions

While the urgency for COVID-19-specific interventions has diminished, antiseptic mouthwashes remain relevant for broader applications in oral and respiratory health. Future research should explore their role in reducing viral loads for other respiratory pathogens, such as influenza, respiratory syncytial virus (RSV), and emerging coronaviruses, which continue to pose public health risks. Additionally, investigating the long-term effects of antiseptic mouthwashes on oral microbiota, periodontal health, gut health, and antimicrobial resistance could provide valuable insights into their broader health benefits. Further studies should also focus on optimizing rinsing protocols, including factors such as rinse duration, frequency, and active ingredient concentrations, to enhance utility.

Conducting larger, well-powered clinical trials with diverse populations and extended follow-up periods would help determine the long-term impact of mouthwashes on viral transmission and overall oral health. Exploring their potential role in reducing bacterial and viral transmission in healthcare settings, schools, and other high-risk environments could also be valuable.

In conclusion, in this pilot randomized clinical trial, we observed no statistically significant differences in SARS-CoV-2 viral load between OTC antiseptic mouthwashes and distilled water. Although some products showed slightly greater increases in Ct values than the control (water) over time, these changes were not clinically meaningful. Throat wash samples demonstrated strong agreement with throat swabs and may offer a reliable alternative when swab collection is not feasible. Given the small sample size, early trial discontinuation, and mixed findings across the literature, larger well-powered studies are needed to determine whether antiseptic mouthwashes have a role in reducing viral transmission.

## Supporting information

CONSORT

Human Participants

## Acknowledgments

We are especially grateful to Brenda Gonzalez Vargas for her substantial support in participant recruitment, delivery of sample collection kits, and retrieval of participants’ collected samples. We sincerely thank Danielle Stephens for initiating the REDCap project and developing the study instruments. We also thank Sarit Helman for her continued support with REDCap management. We gratefully acknowledge Dr. Shakiba Ghasemi Assl for her thoughtful contributions during the manuscript preparation phase. Finally, we thank Mehrdad Yadegari for his assistance with manuscript submission.

## Data Availability

The data underlying the results presented in this study are not publicly available due to ethical and privacy restrictions involving human participants. De-identified data are available from the corresponding author upon reasonable request for researchers who meet the criteria for access to confidential data, subject to approval by the University of California, San Francisco Institutional Review Board (IRB).

## Funding

- University of California Office of the President Emergency COVID-19 Research Funding through the California Breast Cancer Research Program (Grant #R00RG2901)
- NIH/NIDCR T32DE007306 and NIDCR-K23DE033785 (supporting Dr. Banava’s time)
- NIH/NIDCR K23DE033785 (supporting Dr. Banava’s time)
- NIH/NIDCR U01DE025507-S1 (supporting pilot trial coordination)
- CTSI Participant Recruitment Program (PRP) support TL1TR001872
- Rowpar Pharmaceutical Inc. for funding and in-kind product support, and Procter & Gamble for providing in-kind products.

The contents of this report are solely the responsibility of the authors and do not necessarily represent the official views of the NIH or any other funders.

## Author contributions

**Conceptualization:** SB, SG

**Methodology:** SB, SG, AR, PK, YK

**Formal Analysis:** SB, NC, SG

**Data Curation:** SB, AR

**Investigation:** SB, AR

**Laboratory Work:** AR

**Laboratory Supervision:** PK, YK

**Project Administration:** SB

**Participant Recruitment and Sample Collection Coordination:** SB

**Data Visualization:** NC, SB

**Supervision:** SG, SB, YK

**Funding Acquisition:** SG

**Writing – Original Draft:** SB, AR (Parts of Methods section)

**Review & Editing:** All authors

## Competing Interests

Rowpar Pharmaceutical Inc. provided in-kind product support and partial funding for staffing, laboratory assay costs, and supplies. Procter & Gamble provided in-kind product support. The funders had no role in study design, data collection and analysis, decision to publish, or preparation of the manuscript. The authors declare no personal competing interests.

## Notes

### Competing Interest Statement

The authors have declared no competing interest.

### Clinical Trial

ClinicalTrials.gov NCT04409873

### Clinical Protocols

https://clinicaltrials.gov/study/NCT04409873

### Funding Statement

Yes

### Author Declarations

Institutional Review Board of the University of California San Francisco gave ethical approval for this work.

### Summary of Updates

This version of the manuscript has been revised to update the following: 1- Revised the swapped first and last name of the author Pachiyappan Kamarajan (was listed as Kamarajan Pachiyappan). 2- Revised the date formatting as DDMMYYYY. 3- Revised the Data availability. 4- Revised competing interest section.

## References

1. World Health Organization. Number of COVID-19 deaths reported to WHO. Updated 2024. Accessed April 30, 2025. https://data.who.int/dashboards/covid19/deaths

2. Centers for Disease Control and Prevention. COVID Data Tracker. Updated 2024. Accessed February 10, 2025. https://covid.cdc.gov/covid-data-tracker

3. Wu Z, McGoogan JM. Characteristics of and important lessons from the coronavirus disease 2019 (COVID-19) outbreak in China: summary of a report of 72 314 cases from the Chinese Center for Disease Control and Prevention. JAMA. 2020;323(13):1239–1242. doi:10.1001/jama.2020.2648

4. Ramesh S, et al. Emerging SARS-CoV-2 variants: a review of its mutations, its implications and vaccine efficacy. Vaccines (Basel). 2021;9(10):1195. doi:10.3390/vaccines9101195

5. Carabelli AM, et al. SARS-CoV-2 variant biology: immune escape, transmission and fitness. Nat Rev Microbiol. 2023;21(3):162–177. doi:10.1038/s41579-022-00841-7

6. Meng L, Hua F, Bian Z. Coronavirus disease 2019 (COVID-19): emerging and future challenges for dental and oral medicine. J Dent Res. 2020;99(5):481–487. doi:10.1177/0022034520914246

7. Xu J, et al. Salivary glands: potential reservoirs for COVID-19 asymptomatic infection. J Dent Res. 2020;99(8):989. doi:10.1177/0022034520918518

8. da Silva Pedrosa M, et al. Are the salivary glands the key players in spreading COVID-19 asymptomatic infection in dental practice? J Med Virol. 2021;93(1):204–205. doi:10.1002/jmv.26316

9. Bajgain KT, et al. Prevalence of comorbidities among individuals with COVID-19: a rapid review of current literature. Am J Infect Control. 2021;49(2):238–246. doi:10.1016/j.ajic.2020.06.213

10. Magesh S, et al. Disparities in COVID-19 outcomes by race, ethnicity, and socioeconomic status: a systematic review and meta-analysis. JAMA Netw Open. 2021;4(11):e2134147. doi:10.1001/jamanetworkopen.2021.34147

11. Khanijahani A, et al. A systematic review of racial/ethnic and socioeconomic disparities in COVID-19. Int J Equity Health. 2021;20(1):248. doi:10.1186/s12939-021-01582-4

12. Basilicata M, et al. Impact of SARS-CoV-2 on dentistry: a review of literature. Eur Rev Med Pharmacol Sci. 2022;26(9):3386–3398. doi:10.26355/eurrev_202205_28760

13. Characteristics of health care personnel with COVID-19—United States, February 12–April 9, 2020. MMWR Morb Mortal Wkly Rep. 2020;69(15):477–481. doi:10.15585/mmwr.mm6915e6

14. Patel VR, Liu M, Worsham CM, Stanford FC, Ganguli I, Jena AB. Mortality Among US Physicians and Other Health Care Workers. JAMA Intern Med. 2025 May 1;185(5):563–571. doi: 10.1001/jamainternmed.2024.8432. PMID: 39992637; PMCID: PMC11851301

15. Centers for Disease Control and Prevention (CDC). Guidelines for infection control in dental health-care settings—2003. Accessed May 1, 2025. https://www.cdc.gov/mmwr/preview/mmwrhtml/rr5217a1.htm

16. Peng X, et al. Transmission routes of 2019-nCoV and controls in dental practice. Int J Oral Sci. 2020;12(1):9. doi:10.1038/s41368-020-0075-9

17. Marui VC, et al. Efficacy of preprocedural mouthrinses in the reduction of microorganisms in aerosol: a systematic review. J Am Dent Assoc. 2019;150(12):1015–1026.e1. doi:10.1016/j.adaj.2019.06.024

18. Reis INR, et al. Can preprocedural mouthrinses reduce SARS-CoV-2 load in dental aerosols? Med Hypotheses. 2021;146:110436. doi:10.1016/j.mehy.2020.110436

19. Centers for Disease Control and Prevention. Guideline for disinfection and sterilization in healthcare facilities. Updated 2008. Accessed May 1, 2025. https://www.cdc.gov/infection-control/hcp/disinfection-sterilization/chemical-disinfectants.html

20. American Dental Association. Summary of ADA guidance during the COVID-19 crisis. Updated 2020. Accessed May 1, 2025. https://www.ada.org/about/press-releases/2020-archives/summary-of-ada-guidance-during-the-covid-19-crisis

21. Meister TL, et al. Virucidal efficacy of different oral rinses against severe acute respiratory syndrome coronavirus 2. J Infect Dis. 2020;222(8):1289–1292. doi:10.1093/infdis/jiaa471

22. Meister TL, et al. Virucidal activity of a plant-oil–based oral rinse against respiratory viruses. J Hosp Infect. 2024;147:83–86. doi:10.1016/j.jhin.2024.02.023

23. Shewale JG, et al. In vitro antiviral activity of stabilized chlorine dioxide containing oral care products. Oral Dis. 2023;29(3):1333–1340. doi:10.1111/odi.14044

24. Seneviratne CJ, et al. Efficacy of commercial mouth-rinses on SARS-CoV-2 viral load in saliva: a randomized control trial. Infection. 2021;49(2):305–311. doi:10.1007/s15010-020-01563-9

25. Harris PA, et al. Research electronic data capture (REDCap)—a metadata-driven methodology and workflow process for providing translational research informatics support. J Biomed Inform. 2009;42(2):377–381. doi:10.1016/j.jbi.2008.08.010

26. Harris PA, et al. The REDCap consortium: building an international community of software platform partners. J Biomed Inform. 2019;95:103208. doi:10.1016/j.jbi.2019.103208

27. Bidra AS, et al. Rapid in vitro inactivation of severe acute respiratory syndrome coronavirus 2 (SARS-CoV-2) using povidone-iodine oral antiseptic rinse. J Prosthodont. 2020;29(6):529–533. doi:10.1111/jopr.13209

28. Engelmann I, et al. Preanalytical issues and cycle threshold values in SARS-CoV-2 real-time RT-PCR testing: should test results include these? ACS Omega. 2021;6(10):6528–6536. doi:10.1021/acsomega.1c00166

29. Badu K, et al. SARS-CoV-2 viral shedding and transmission dynamics: implications of WHO COVID-19 discharge guidelines. Front Med (Lausanne). 2021;8:648660. doi:10.3389/fmed.2021.648660

30. Onozuka D, et al. Oral mouthwashes for asymptomatic to mildly symptomatic adults with COVID-19 and salivary viral load: a randomized, placebo-controlled, open-label clinical trial. BMC Oral Health. 2024;24(1):491. doi:10.1186/s12903-024-04246-1

31. Ferrer MD, et al. Clinical evaluation of antiseptic mouth rinses to reduce salivary load of SARS-CoV-2. Sci Rep. 2021;11(1):24392. doi:10.1038/s41598-021-03461-y

32. Mezarina Mendoza JPI, et al. Antiviral effect of mouthwashes against SARS-CoV-2: a systematic review. Saudi Dent J. 2022;34(3):167–193. doi:10.1016/j.sdentj.2022.01.006

33. Tarragó-Gil R, et al. Randomized clinical trial to assess the impact of oral intervention with cetylpyridinium chloride to reduce salivary SARS-CoV-2 viral load. J Clin Periodontol. 2023;50(3):288–294. doi:10.1111/jcpe.13746

34. Alemany A, et al. Cetylpyridinium chloride mouthwash to reduce shedding of infectious SARS-CoV-2: a double-blind randomized clinical trial. J Dent Res. 2022;101(12):1450–1456. doi:10.1177/00220345221102310

35. Bezinelli LM, et al. Reduction of SARS-CoV-2 viral load in saliva after rinsing with mouthwashes containing cetylpyridinium chloride. PeerJ. 2023;11:e15080. doi:10.7717/peerj.15080

36. Alzahrani MM, et al. Mouth rinses efficacy on salivary SARS-CoV-2 viral load: a randomized clinical trial. J Med Virol. 2023;95(1):e28412. doi:10.1002/jmv.28412

37. Perussolo J, et al. Efficacy of three antimicrobial mouthwashes in reducing SARS-CoV-2 viral load in the saliva of hospitalized patients: a randomized controlled pilot study. Sci Rep. 2023;13(1):12647. doi:10.1038/s41598-023-39308-x

38. Azzi L, et al. Saliva is a reliable tool to detect SARS-CoV-2. J Infect. 2020;81(1):e45–e50. doi:10.1016/j.jinf.2020.04.005

39. Iwasaki S, et al. Comparison of SARS-CoV-2 detection in nasopharyngeal swab and saliva. J Infect. 2020;81(2):e145–e147. doi:10.1016/j.jinf.2020.05.071

40. Zhu J, et al. Viral dynamics of SARS-CoV-2 in saliva from infected patients. J Infect. 2020;81(3):e48–e50. doi:10.1016/j.jinf.2020.06.059

41. Sakanashi D, et al. Comparative evaluation of nasopharyngeal swab and saliva specimens for molecular detection of SARS-CoV-2 RNA. J Infect Chemother. 2021;27(1):126–129. doi:10.1016/j.jiac.2020.09.027

42. Wyllie AL, et al. Saliva or nasopharyngeal swab specimens for detection of SARS-CoV-2. N Engl J Med. 2020;383(13):1283–1286. doi:10.1056/NEJMc2016359

43. Centers for Disease Control and Prevention. Symptoms of COVID-19. Updated 2025. Accessed May 1, 2025. https://www.cdc.gov/covid/signs-symptoms/index.html

44. Karami H, et al. Comparison of the effects of chlorhexidine and sodium bicarbonate mouthwashes on COVID-19–related symptoms. Iran J Nurs Midwifery Res. 2024;29(1):60–67. doi:10.4103/ijnmr.ijnmr_38_23

45. Poleti ML, et al. Use of mouthwash and dentifrice containing an antimicrobial phthalocyanine derivative for the reduction of clinical symptoms of COVID-19: a randomized triple-blind clinical trial. J Evid Based Dent Pract. 2022;22(4):101777. doi:10.1016/j.jebdp.2022.101777

46. Brookes Z, et al. Mouthwash effects on the oral microbiome: are they good, bad, or balanced? Int Dent J. 2023;73(suppl 2):S74–S81. doi:10.1016/j.identj.2023.08.010

47. Schwarz SR, et al. Limited antimicrobial efficacy of oral care antiseptics in microcosm biofilms and phenotypic adaptation of bacteria upon repeated exposure. Clin Oral Investig. 2021;25(5):2939–2950. doi:10.1007/s00784-020-03613-w

48. Cavalcante-Leão BL, et al. Is there scientific evidence of the mouthwashes effectiveness in reducing viral load in COVID-19? A systematic review. J Clin Exp Dent. 2021;13(2):e179–e189. doi:10.4317/jced.57406

49. Vecchio J, et al. Viral and immunologic evaluation of smokers with severe COVID-19. Sci Rep. 2023;13(1):17898. doi:10.1038/s41598-023-45195-z

50. Gregorczyk-Maga I, et al. Impact of tooth brushing on oral bacteriota and health care-associated infections among ventilated COVID-19 patients: an intervention study. Antimicrob Resist Infect Control. 2023;12(1):17. doi:10.1186/s13756-023-01218-y

51. Natto ZS, et al. Short-term effect of different chlorhexidine forms versus povidone iodine mouth rinse in minimizing the oral SARS-CoV-2 viral load. Medicine (Baltimore). 2022;101(30):e28925. doi:10.1097/MD.0000000000028925

