## Appendix for "Effect of Antiseptic Mouthwash/Gargling Solutions on SARS-CoV-2 Viral Load: A Randomized Clinical Trial"

### Effect of Antiseptic Mouthwash/Gargling Solutions and Pre-Procedural Rinse on SARS-CoV-2 Load: A Randomized Clinical Trial

#### Supplemental Materials

#### Appendix A – Least Squares Means analysis

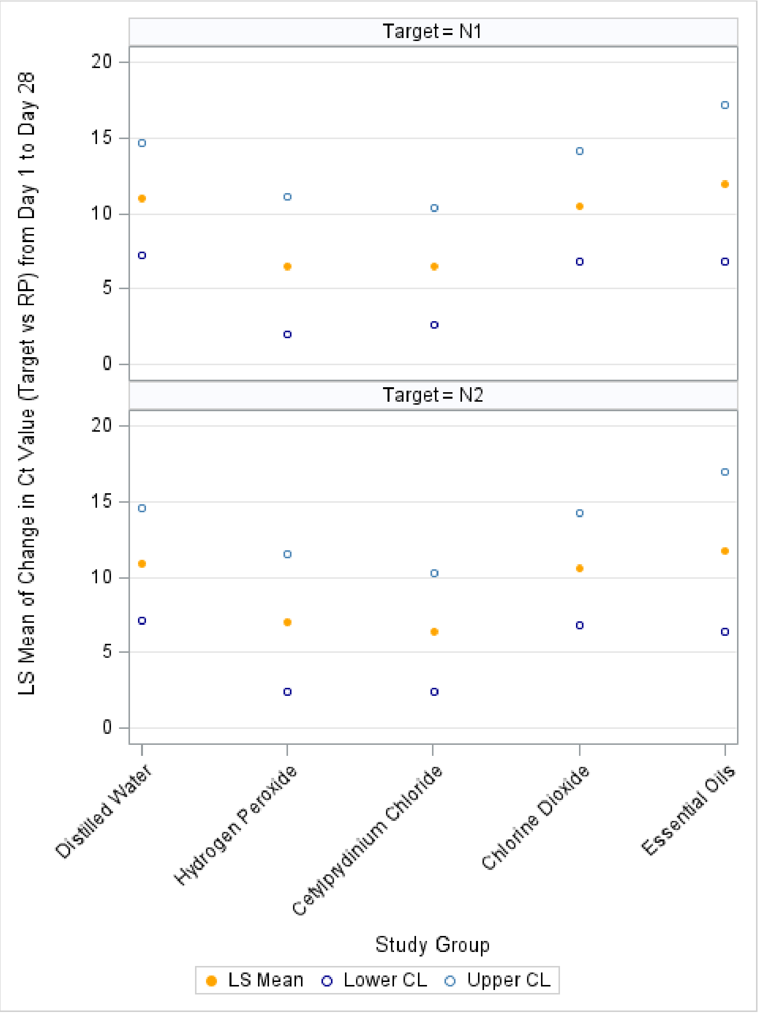


**Least squares mean change in Ct values from Day 1 to Day 28 by study group.** Plots display the least squares (LS) mean change in Ct values (target Ct normalized to RP) from Day 1 to Day 28 for the N1 target and N2 target. Points represent LS mean estimates, with corresponding lower and upper 95% confidence limits shown as separate markers. Positive values indicate an increase in Ct (corresponding to lower detectable viral RNA over time). Adjusted mean Ct values increased from Day 1 to Day 28 across all study groups for both N1 and N2 targets, indicating reduced detectable viral RNA over time. Essential Oils and Chlorine Dioxide show the largest adjusted mean increases, while Hydrogen Peroxide and Cetylpyridinium Chloride demonstrate smaller changes. The proximity of confidence limits across groups suggests overlapping estimates and no clear evidence of a distinct treatment effect separating one group from the others.

**Appendix B- Baseline Questionnaire**

BaselineQuestionnaireCuestiona

**Appendix C – Sample Collection Procedure**

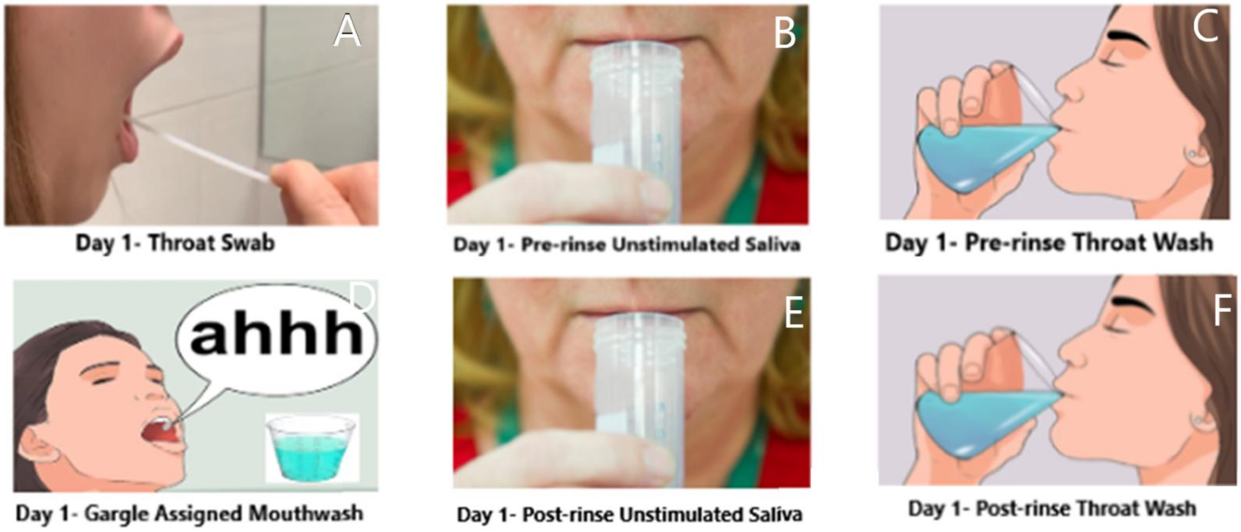

#### Appendix D- Participant History of Tobacco, Marijuana, and Related Product Use Across Study Groups

**Appendix Table 1 - Participant History of Tobacco, Marijuana, and Related Product Use Across Study Groups**

| | Distilled Water (N=12) | Hydrogen Peroxide (N=11) | Cetylpyridinium Chloride (N=12) | Chlorine Dioxide (N=12) | Essential Oils (N=7) | Total (N=54) |
|---|---|---|---|---|---|---|
| **Ever used Tobacco products*** | | | | | | |
| Cigarettes | 5 (42) | 6 (55) | 3 (25) | 5 (42) | 3 (43) | 22 (41) |
| E-cigarettes or other electronic nicotine products | 3 (25) | 2 (18) | 2 (17) | 2 (17) | 0 (0) | 9 (17) |
| Marijuana, THC, or CBD | 6 (50) | 6 (55) | 1 (8) | 7 (58) | 3 (43) | 23 (43) |
| Traditional cigars | 3 (25) | 0 (0) | 1 (8) | 2 (17) | 0 (0) | 6 (11) |
| Shisha or hookah tobacco | 2 (17) | 4 (36) | 0 (0) | 2 (17) | 1 (14) | 9 (17) |
| Other types of smokeless tobacco | 1 (8) | 0 (0) | 0 (0) | 0 (0) | 0 (0) | 1 (2) |
| None of them | 5 (42) | 3 (27) | 6 (50) | 4 (33) | 3 (43) | 21 (39) |
| **Smoked a cigarette in the past 12 months*** | | | | | | |
| Yes | 4 (33) | 0 (0) | 1 (8) | 0 (0) | 0 (0) | 5 (9) |
| No | 8 (67) | 11 (100) | 11 (92) | 12(100) | 7 (100) | 49 (91) |
| **Smoked a cigarette in the past 30 days*** | | | | | | |
| Yes | 2 (17) | 0 (0) | 1 (8) | 0 (0) | 0 (0) | 3 (6) |
| No | 10 (83) | 11 (100) | 11 (92) | 12(100) | 7 (100) | 51 (94) |
| **Smoked marijuana/THC/CBD in the past 12 months**** | | | | | | |
| Yes | 3 (25) | 4 (36) | 1 (8) | 5 (42) | 2 (29) | 15 (28) |
| No | 9 (75) | 7 (64) | 11 (92) | 7 (58) | 5 (71) | 39 (72) |
| **Smoked marijuana/THC/CBD in the past 30 days**** | | | | | | |
| Yes | 2 (17) | 3 (27) | 0 (0) | 4 (33) | 2 (29) | 11 (20) |
| No | 10 (83) | 8 (73) | 12 (100) | 8 (67) | 5 (71) | 43 (80) |
| **Use any tobacco product soon after waking** | | | | | | |
| **0-60 minutes***** | 0 (0) | 0 (0) | 0 (0) | 0 (0) | 0 (0) | 0 (0) |
| **More than 60 minutes** | 3 (100) | 2 (100) | 1 (100) | 2 (100) | 1 (50) | 9 (90) |
| **Prefer not to answer** | 0 (0) | 0 (0) | 0 (0) | 0 (0) | 1 (50) | 1 (10) |

Note: * Even one or two puffs/tries; ** even one or two times; *** This category combines the groups “within 5 minutes,” “6–30 minutes,” and “31–60 minutes,” as there were no participants in these categories.

#### Appendix E- Participant-Reported Symptoms and Feedback Related to Mouthwash Use

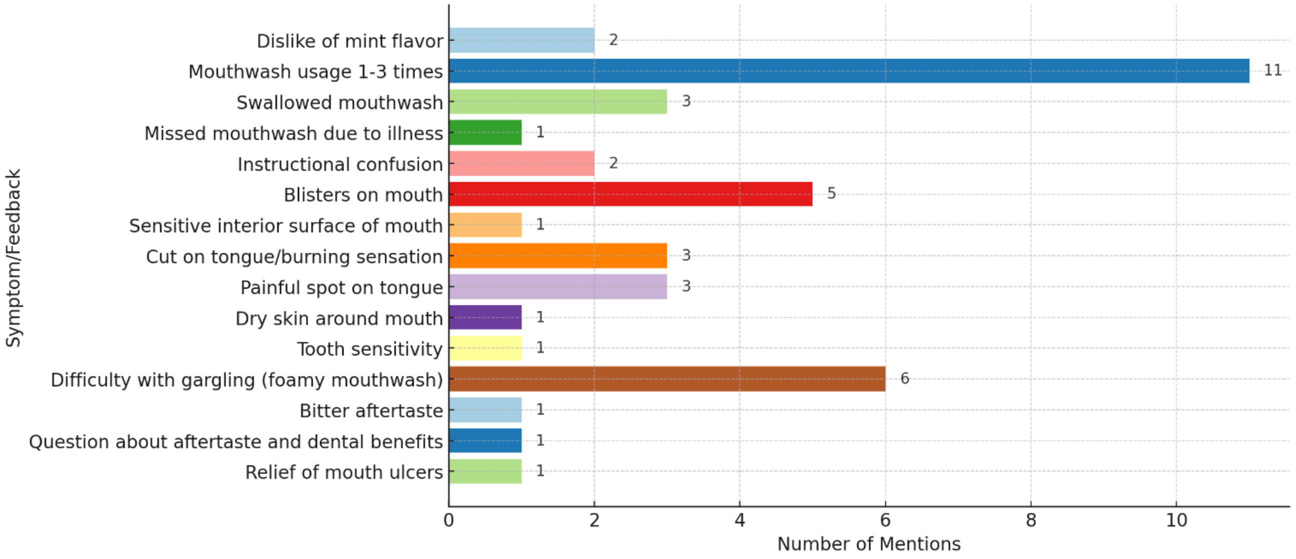


Horizontal bar chart displays the number of mentions for self-reported symptoms, adverse effects, and general feedback associated with mouthwash use during the study period. The x-axis represents the number of mentions, and the y-axis lists reported symptoms or feedback categories. The most frequently reported feedback was study mouthwash use of 1–3 times (n=11), reflecting adherence-related reporting. Among adverse effects, difficulty with gargling (foamy mouthwash) (n=6) and blisters in the mouth (n=5) were the most commonly mentioned. Reports of swallowed mouthwash (n=3), cut on the tongue or burning sensation (n=3), and painful spot on the tongue (n=3) were also noted.

Less frequently reported issues included dislike of mint flavor (n=2), instructional confusion (n=2), and single mentions of sensitive oral mucosa, dry skin around the mouth, tooth sensitivity, bitter aftertaste, questions about aftertaste and dental benefits, missed use due to illness, and perceived relief of mouth ulcers. In general, reported symptoms were infrequent and varied, with no single adverse effect predominating beyond mild local oral discomfort.

#### Appendix F- Oral Hygiene Practices among Study Participants at Baseline (Day 1).

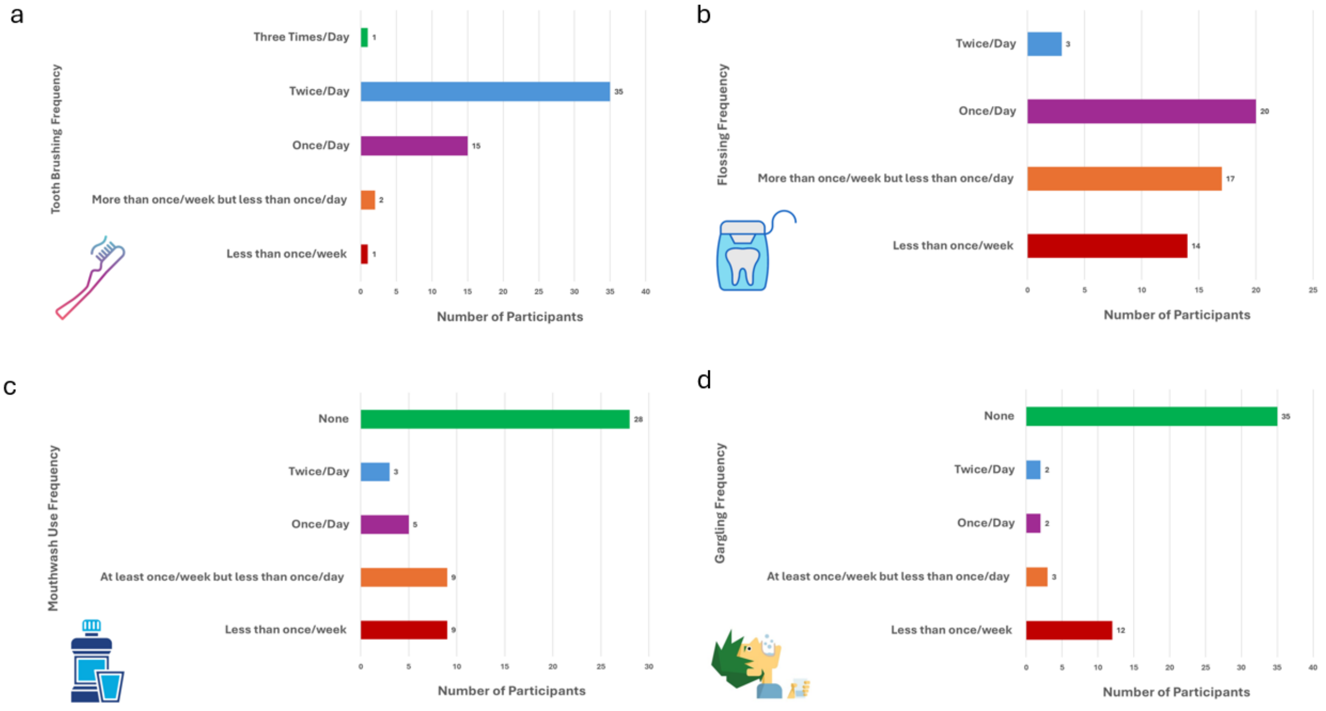


Bar charts display self-reported frequencies of (a) tooth brushing, (b) flossing, (c) mouthwash use, and (d) gargling among study participants. The x-axis represents the number of participants, and the y-axis lists frequency categories for each behavior.

Most participants reported brushing their teeth twice daily, with fewer individuals brushing once daily or more than twice per day. In contrast, flossing was less frequent overall, with the largest proportions reporting once daily flossing or flossing more than once per week but less than daily; a notable proportion reported flossing less than once per week. Mouthwash use was infrequent for many participants, with the largest group reporting no use. Similarly, gargling was uncommon, with the majority reporting no gargling and relatively few participants engaging in daily or twice-daily gargling.
