## Supplementary material for "Effect of Antiseptic Mouthwash/Gargling Solutions on SARS-CoV-2 Viral Load: A Randomized Clinical Trial": Human Participants

### Human Participants Research Checklist

#### Complete the following if your study involved human participants or human participants' data. These questions should be addressed for prospective and retrospective studies.

1. Did you obtain ethics approval for this study?

- If yes, please upload all the approval documents you received from your ethics committee to cover the entire range of the study period (file type "Other").
    - Please include the original approval document and any extension documents
    - Where ethics approval was obtained from more than one study location, please provide approval document(s) from all of the sites.
    - If the original document is in another language, please also provide an English translation.

X Uploaded ___ N/A

- If either of the following applies to your submission, please explain the reasons in the space below:
    - You did not obtain ethical approval.
    - You performed an international study, but did not obtain ethical approval from all countries where data collection took place.

Continues on page 2.

2. If you prospectively recruited human participants for the study – for example, you conducted a clinical trial, distributed questionnaires, or obtained tissues, data or samples for the purposes of this study, please report in the Methods:

    i. the day, month and year of the **start and end** of the recruitment period for this study.

    ii. whether participants provided informed consent, and if so, what type was obtained (for instance, written or verbal, and if verbal, how it was documented and witnessed). If your study included minors, state whether you obtained consent from parents or guardians. If the need for consent was waived by the ethics committee, please include this information.

   Please state the line number(s) in the Methods where this is reported ______

   ___ Completed   _X_ N/A

3. If you are reporting a retrospective study of, for example, medical records, archived samples, survey data, please report in the Methods section:

    ii. the day, month and year when the data were accessed for research purposes

    iii. whether authors had access to information that could identify individual participants during or after data collection.

   Please state the line number(s) in the Methods where this is reported ______

   ___ Completed   _X_ N/A
